# Accurate cross-platform GWAS analysis via two-stage imputation

**DOI:** 10.1101/2024.04.19.24306081

**Authors:** Anya Greenberg, Kaylia M. Reynolds, Zhanqing Hua, Michelle T. McNulty, Matthew G. Sampson, Hyun M. Kang, Dongwon Lee

## Abstract

In genome-wide association studies (GWAS), combining independent case-control cohorts has been successful in increasing power for meta and joint analyses. This success sparked interest in extending this strategy to GWAS of rare and common diseases using existing cases and external controls. However, heterogeneous genotyping data can cause spurious results. To harmonize data, we propose a new method, two-stage imputation (TSIM), where cohorts are imputed separately, merged on intersecting high-quality variants, and imputed again. We show that TSIM controls imputation-derived errors and type I error. Merging arthritis cases and UK Biobank controls using TSIM, we replicated known associations without introducing false positives. Furthermore, GWAS using TSIM performed comparably to the meta-analysis of nephrotic syndrome cohorts genotyped on five different platforms, demonstrating TSIM’s ability to harmonize heterogeneous genotyping data. With the plethora of publicly available genotypes, TSIM provides a GWAS framework that harmonizes heterogeneous data, enabling analysis of small and case-only cohorts.

## Introduction

Genome-wide association studies (GWAS) are common for studying the genetic determinants of traits and diseases, especially when large sample sizes are available. Smaller studies or case-only cohorts are often combined with external controls to enable GWAS and increase statistical power^1–3^. When combining cohorts, batch effects can arise due to differences in genotyping platform. To reduce these effects, the current practice for combining cohorts prior to imputation is to use genotyped single nucleotide polymorphisms (SNPs) that are shared between cohorts^1,4–7^. This approach not only reduces the number of SNPs available for analysis but may also adversely affect genotype imputation by decreasing accuracy.

Genotype imputation approximates whole genome sequencing (WGS) by leveraging linkage disequilibrium (LD)^8,9^. It is an essential step in GWAS to increase the genome-wide coverage of the analysis and enable post-GWAS analyses such as fine mapping and meta-analysis^1,9^. By restricting the SNPs used for imputation to the intersection, haplotype-informative SNPs may be lost, thus reducing the accuracy of imputation and downstream analyses. Given this, it is currently recommended to only combine cohorts genotyped on the same platform prior to imputation^4,5^. Finding external cohorts that meet these criteria is often difficult, especially for underrepresented populations. While it is becoming easier to find large external control cohorts with the advent of diverse biobanks, they are often genotyped on custom platforms designed for their population of interest^10,11^ making it difficult to combine them with other cohorts.

Previously, several approaches have been proposed to overcome the challenges in combining heterogeneous genotyping data. Some studies argued that cohorts can be imputed separately, and then combined by restricting analysis to the shared SNPs with high imputation quality^2,12^. This approach reduced type I error in GWAS at the expense of reduced power and fewer SNPs for testing. GAWMerge presented another solution to this problem: merge the array-based genotypes of a study cohort with the WGS of publicly available controls^13^. However, this method requires the external cohort to have WGS available which may not be feasible for many underrepresented populations, particularly in regions with limited funding and resources. Another study sought to harmonize multiple control cohorts by removing low-quality genotyped SNPs, then merging genotypes after a single imputation^7^. These SNPs were identified through an iterative process of GWAS using genotyping platform as the outcome, to identify spurious genome-wide significant loci, and re-imputation on array genotypes, to identify low-quality SNPs. This approach showed promise in reducing batch effects when merging multiple cohorts. However, it requires internal controls genotyped on the same platform as cases. It may also result in substantial loss of imputed SNPs after filtering out low-quality variants from re-imputation.

Here, we introduce two-stage imputation (TSIM), a method to address these issues, which includes a second imputation on high-quality SNPs after merging separately imputed cohorts. In brief, cohorts are imputed separately (Stage 1) and then merged on high-quality genotyped and imputed SNPs (hq-SNPs) present in both imputations. Finally, a second imputation (Stage 2) is performed on the merged data. Essentially, TSIM utilizes a first stage of high-quality imputation to improve a second stage of imputation which covers a larger proportion of the genome. Similar strategies have been used to improve genotype imputation of ancient DNA^14^ and rare variants^15^, but no studies have applied this concept to GWAS of common variants. We illustrate the validity and utility of TSIM using data from UK Biobank (UKB)^10^, a psoriatic arthritis case-control cohort^16^, and two cohorts from a published pediatric steroid sensitive nephrotic syndrome (pSSNS) meta-analysis^17^. Along with this research, we provide a command line tool, <u>tsim</u>, for others to easily implement this method in their analyses.

## Results

### Overview of two-stage imputation method (TSIM)

TSIM implements two primary steps bookended by two rounds of imputation (**Fig. 1**). In the first round of imputation (Stage 1), we impute cohorts separately using the same reference panel, following current GWAS practices for quality control (QC) of a single cohort (**Methods**). This Stage 1 imputation serves to increase the number of intersecting SNPs for merging by identifying high-quality genotyped and imputed SNPs (hq-SNPs) present in all imputed cohorts. We define hq-SNPs as common SNPs (minor allele frequency (MAF) ≥ 0.01) with high imputation quality (R^2^ ≥ 0.98; empirical R^2^ [ER^2^] ≥ 0.9 if genotyped). We discuss the determination of these thresholds in detail below. This step can be considered as *“in silico* genotyping*”* simulating the “same platform” for all cohorts to be merged. After Stage 1, cohorts are merged on the intersection of hq-SNPs. Then, we perform the second round of imputation (Stage 2) on the merged cohort to achieve greater genome-wide coverage for GWAS and downstream analyses.

**Figure 1:**
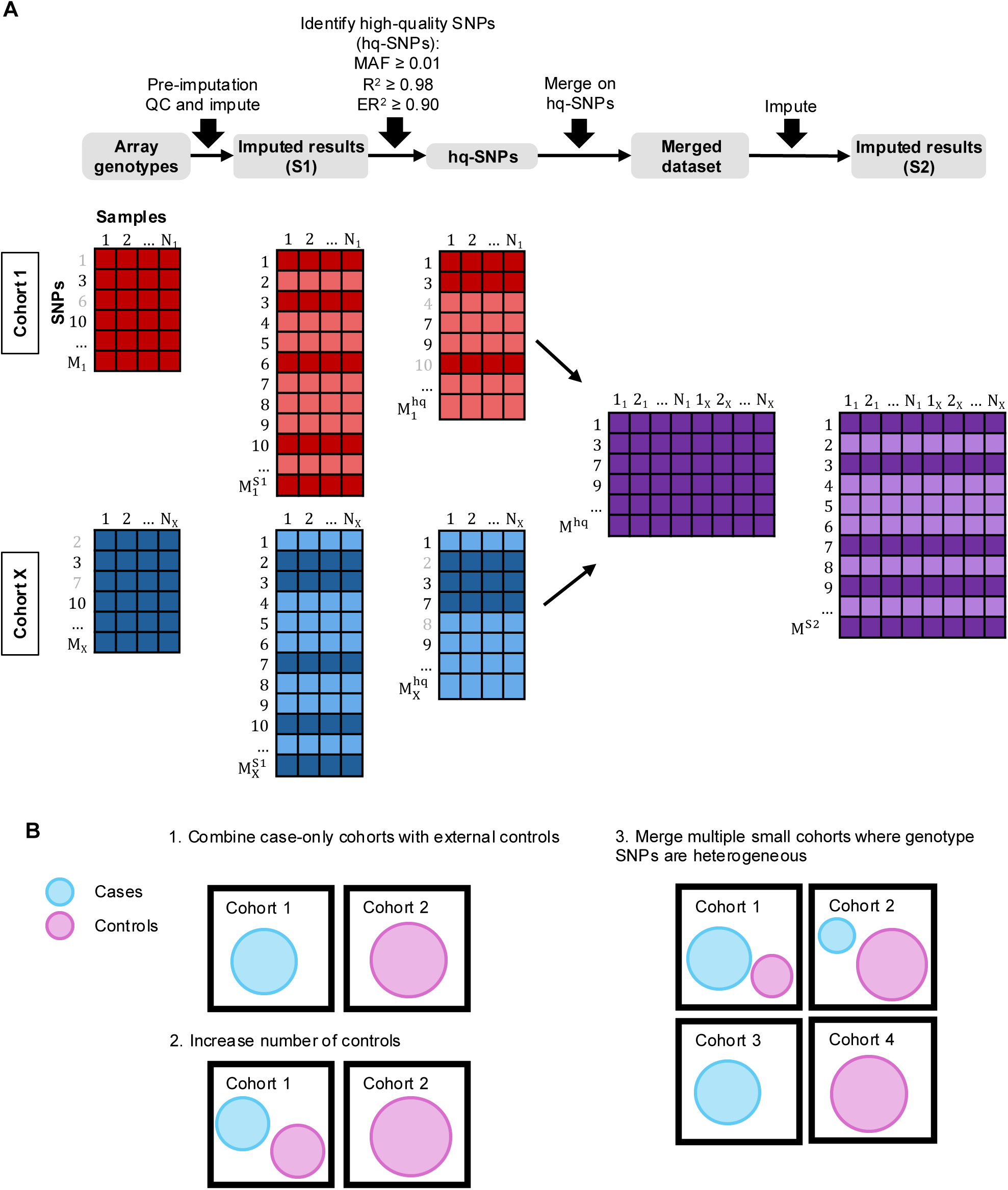
An overview of the two-stage imputation method and its applications. **(A)** A detailed workflow for the two-stage imputation method applied to X number of cohorts. Dark shaded regions represent SNPs included in input to imputation. Light shaded regions represent imputed SNPs. **(B)** Illustrations of potential applications of TSIM.

### Determination of high-quality SNPs

We evaluated the quality of Stage 1 imputation results to determine what criteria were needed to define our hq-SNPs. We used unrelated individuals from the UK Biobank (UKB) across five ancestry groups (European [EUR], African [AFR], Admixed American [AMR], East Asian [EAS], and South Asian [SAS]) who were genotyped on the UKB Axiom array and had whole genome/exome sequencing (WGS/WES) data. We imputed genotypes using the TOPMed Imputation Server and its associated reference panel (TOPMed)^18–20^. Within each ancestry group, we identified imputation-derived errors based on the overall correlation between Stage 1 imputed genotypes and the WGS genotypes, which we considered the ground truth. Specifically, we first split the SNPs by their imputation quality (measured by R^2^) into five groups with increments of 0.01 from 0.95 to 1.00. We then calculated correlation values (r^2^) for each group of SNPs, further stratified by their minor allele frequencies (MAF) using GLIMPSE2^21,22^ (**Methods**). In this analysis, we evaluated imputed SNPs only, as directly genotyped SNPs have distinct properties, as shown below. As a baseline, we calculated the correlation between WGS and the unimputed genotypes. We found that imputed SNPs with R^2^ ≥ 0.98 generally had r^2^ values more similar to the unimputed baseline across all ancestry groups. For SNPs with R^2^ in the [0.98,0.99) bin, the correlations were 0.97 or higher and were within 0.03 of the respective baselines for each ancestry group. While SNPs with R^2^ ≥ 0.99 had higher correlations, including SNPs in the [0.98,0.99) R^2^ bin doubled the number of SNPs for EAS and quadrupled for EUR, AFR, AMR, and SAS. We also found that rare SNPs tended to perform worse compared to the baseline. For all ancestry groups, the SNPs in the [0.01,0.02) MAF bin had consistently lower correlations with WGS compared to SNPs with higher MAFs. The other R^2^ bins followed in descending order, indicating that SNPs with higher R^2^ had higher concordance with WGS (**Fig. 2A; Supplemental Table S1**).

**Figure 2:**
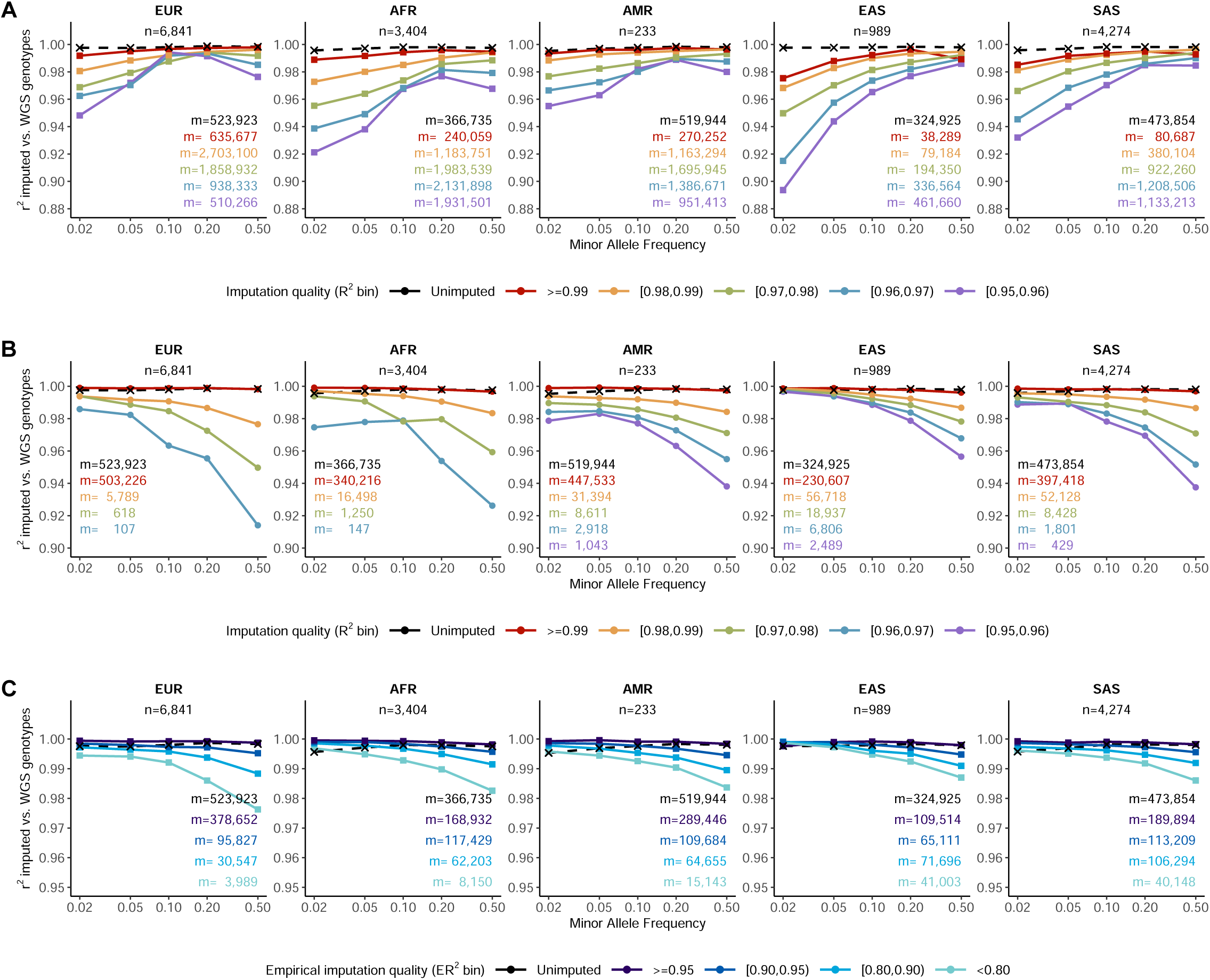
Determination of high-quality SNPs. Line plots show correlation between imputed and WGS genotypes as a function of MAF after the first stage of imputation stratified by whether the SNPs were **(A)** imputed-only, **(B)** present on the genotyping array, or **(C)** present on the genotyping array with R^2^ ≥ 0.98 for UKB Europeans (EUR), Africans (AFR), Admixed Americans (AMR), East Asians (EAS), and South Asians (SAS). SNPs with R^2^ ≥ 0.95 were included, which were then stratified by imputation quality into R^2^ bins (A-B) or empirical R^2^ (ER^2^) bin (C). Unimputed (with ER^2^ ≥ 0.9) vs. WGS (dotted line) shows correlation between unimputed SNP genotypes compared to WGS as a baseline for comparison. The total number of samples is *n* (top center) and the total number of SNPs in each stratum is *m* and colored by R^2^ bins (A-B) or ER^2^ bins (C) (lower right). The y-axis shows the correlation and the x-axis shows the MAF.

Next, we identified the subset of imputed SNPs that were also directly genotyped on the UKB Axiom array and evaluated them separately to better inform our R^2^ cutoff and empirical imputation quality cutoff (ER^2^) for our hq-SNPs. We found that imputed genotypes of these SNPs with R^2^ ≥ 0.98 and ER^2^ ≥ 0.90 had the highest correlation with WGS genotypes and, similarly, were better matched with the baseline (unimputed vs. WGS line) across all ancestry groups (**Fig. 2B,C; Supplemental Table S2,S3**). We note that we used imputed genotypes even at positions that were directly genotyped, to make inputs comparable across platforms. For SNPs in the [0.98,0.99) R^2^ bin, the correlations with WGS were 0.98 or higher and were within 0.02 of the respective baseline for each ancestry group. Furthermore, the majority of SNPs fell into the R^2^ ≥ 0.99 bin which had r^2^ values exceeding the baseline in some cases, i.e., AFR, AMR, and SAS at MAF in [0.01,0.05). Additionally, we found that common SNPs (MAF ≥ 0.05) tended to have lower r^2^ values compared to rare SNPs in the [0.01, 0.05) MAF bin.

Using a similar framework, but with WES genotypes acting as ground truth, we found similar results showing that R^2^ ≥ 0.98 and ER^2^ ≥ 0.90 were sufficient cutoff points for defining our hq-SNPs to be used in the second stage of imputation (**Supplemental Fig. S1; Supplemental Table S4-S6)**. We note that thresholds for R^2^ and ER^2^ should be adjusted accordingly depending on the reference panels, imputation tools, and ancestry groups, although we believe that the criteria established here should work broadly.

### TSIM is robust against imputation-derived errors

Using the hq-SNPs established in the previous step, we followed our TSIM method and performed the Stage 2 imputation on the UKB EUR and the control group (n=1,406) from the psoriatic arthritis (ART) cohort, which is genotyped on the HumanOmni1-Quad BeadChip^16^. Using the UKB EUR dataset, we assessed the effect of TSIM on imputation-derived error by comparing imputed genotypes between Stage 1 and Stage 2. Similar to previous analyses, concordance between imputed and WGS genotypes was calculated as a correlation using GLIMPSE2^21,22^, but we extended the previous analysis to include lower R^2^ bins, as typical GWAS studies include SNPs with R^2^ as low as 0.3. We note that we only assessed UKB EUR individuals due to the limited availability of matching cohorts for our other datasets. Here, we use “S1” and “S2” to represent imputation results following Stage 1 and Stage 2, respectively. S1 is more similar to current practices of imputing once, whereas S2 implements TSIM and imputes data twice. Across R^2^ bins, S1 and S2 demonstrate similar concordance with WGS (**Fig. 3**; **Supplemental Fig. S2; Supplemental Table S7**). The majority of imputed variants fall within the [0.95,1.00] R^2^ bin, indicating high imputation quality. In this bin, the difference in r^2^ between S1 and S2 is < 0.01 for all MAF bins; the maximum difference of 0.0076 (0.9793 vs. 0.9716) was observed in the [0.01, 0.02) MAF bin. While there are more variants with lower R^2^ in S2, the absolute r^2^ difference between S1 and S2 is consistently < 0.06 when all R^2^ bins are considered. The differences tend to be larger in the lower-quality R^2^ bins, without a consistent pattern across MAF bins. Notably, in certain combinations, S2 achieved a better r^2^ than S1: in the [0.30,0.50) R^2^ bin and the [0.02,0.05) MAF bin, S2 outperforms S1 by 0.035 (0.9600 vs. 0.9249), and in the [0.50,0.70) R^2^ bin and the [0.05,0.10) MAF bin, by 0.025. However, these differences may reflect noisy estimation, as the lower-quality R2 bins contain very few variants, especially in S1. We also conducted similar analyses using WES genotypes as the ground truth and found consistent results (**Supplemental Fig. S3; Supplemental Table S8**). Thus, despite using a second stage of imputation where new errors might be introduced, we observed a similar imputation error rate when using TSIM, provided that lower-quality variants are excluded and the reference panel is well-matched to the target.

**Figure 3:**
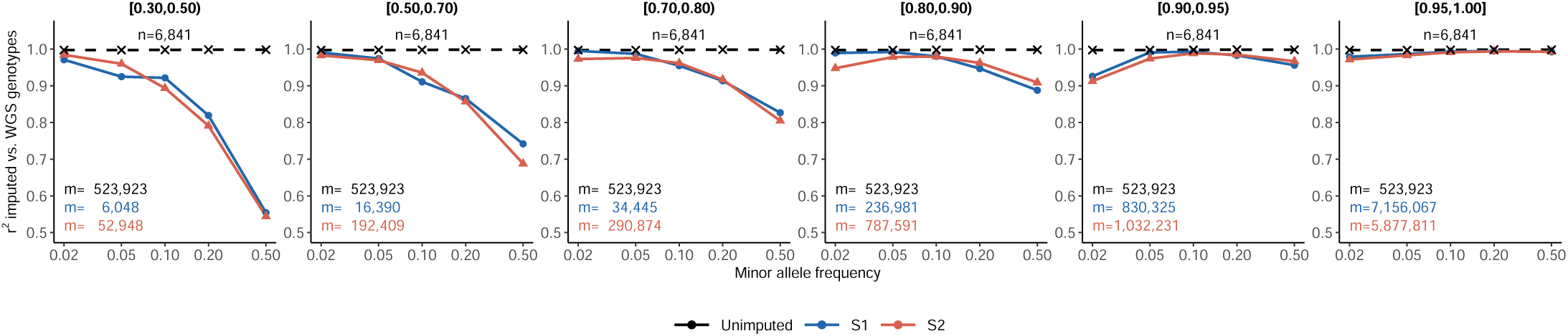
TSIM shows comparable imputation-derived error for S1 and S2. Line plots show correlation between imputed and WGS genotyped as a function of MAF after the first (S1, blue) and second stages (S2, red) of imputation for UKB Europeans. SNPs with R^2^ ≥ 0.30 were included, which were then stratified by imputation quality into R^2^ bins. Unimputed (with ER^2^ ≥ 0.9) vs. WGS (dotted line) shows correlation between unimputed SNP genotypes compared to WGS for comparison. The total number of SNPs in each stratum is *m* and colored by R^2^ bins (lower left). The y-axis shows the correlation and the x-axis shows the MAF.

### TSIM robustly controls for type I error

To evaluate type I error in GWAS, we next performed a GWAS of UKB samples vs. the control group from the ART cohort described above^16^. We used a random subset of 5,000 UKB individuals (UKB5K) of EUR descent, from which individuals with psoriatic arthritis were excluded (**Methods**). Because this was a control-versus-control GWAS, any significant association observed is likely a false positive (type I error). ART controls were merged with UKB5K using TSIM. As a control experiment, we also performed a *separately-imputed* GWAS, in which cohorts were merged after single, separate imputations. In the *TSIM* GWAS, no variants reached genome-wide significance, while the *separately-imputed* GWAS yielded 111 GWAS loci (**Supplemental Fig. S4**). This drastic decrease in GWAS loci between the *separately-imputed* and *TSIM* GWAS, and the noticeable lack of associations in *TSIM,* demonstrates that our method effectively controls for type I error and batch effects across different genotyping platforms, provided there is sufficient genome-wide coverage of overlapping hq-SNPs.

### TSIM replicates psoriatic arthritis GWAS results and enables the use of case-only cohorts

Using the ART cohort, which contained 1,410 psoriatic arthritis cases in addition to the 1,406 controls, and UKB5K, we also evaluated TSIM’s ability to accurately identify GWAS loci in a case-control study. We compared GWAS results from *TSIM* and *separately-imputed* for two different scenarios: (1) *both-controls*: ART cases and controls merged with UKB5K to represent a common situation where external controls may be used to increase the power of GWAS and (2) *external-controls*: only cases from ART merged with UKB5K to represent a situation where only cases were available and merging with external cohorts would enable GWAS to be performed. We also ran GWAS using only the ART cases and controls, referred to as *Published*, to generate reference results to better compare GWAS loci (**Methods**). PCA, QQ plots, and metadata for these GWAS are available in **Supplemental Fig. S4, S5, and Supplemental Table S9**. We found that for both *both-controls* and *external-controls* scenarios, the TSIM GWAS was better able to replicate the *Published* GWAS than the *separately-imputed* GWAS (**Fig. 5A**; **Supplemental Fig. S6**). Specifically, in the *both-controls* GWAS, three out of four previously published genome-wide significant loci from the *Published* GWAS had similar λ_GC_-adjusted p-values, odds ratios, and directions of effect (*HLA-B* – rs13208617; *TRAF3IP2* – rs33980500; *IL12B* – rs918520, rs4921483; *TRAF3IP2* – rs33980500) with the fourth reaching suggestive significance (*TYK2* –rs35251378 [p_λGC-adj._=4.93×10^-7^]) (**Supplemental Fig. S7**, **Supplemental Table S10**). The *both-controls* GWAS also replicated and improved results for some loci by bringing them more confidently over the genome-wide significance threshold (*HLA-G* – rs3115628 (p_λGC-adj._=1.90×10^-9^); *TNIP1* – rs75851973 (p_λGC-adj._=4.92×10^-12^), rs17728338 (p_λGC-adj._=1.65×10^-11^)) (**Fig. 5B**; **Supplemental Fig. S7**, **Supplemental Table S10**). These suggestive loci were implicated in previously published psoriatic arthritis GWAS and many had follow-up analyses further connecting the loci to the disease risk^16,23–26^. Similarly, the *external-controls* GWAS replicated three out of four signals from the *Published* GWAS (*HLA-B* – rs13208617; *TRAF3IP2* – rs33980500; *IL12B* – rs4921483). We also found improved results for one suggestive loci (*TNIP1* – rs75851973 (p_λGC-adj._=4.18×10^-10^), rs17728338 (p_λGC-adj._=1.70×10^-10^)) from the *Published* GWAS (**Fig. 5B**; **Supplemental Fig. S7; Supplemental Table S10**). On the other hand, *separately-imputed* resulted in many false positives, similar to the previous ART control vs. UKB control GWAS (**Fig. 4**). These GWAS demonstrate that our TSIM method can accurately replicate and improve GWAS results by enabling the inclusion of external controls to increase the power of detection. It is especially effective for case-only cohorts for which GWAS analysis was previously impossible due to a lack of controls matching both genetic population structure and genotyping platform.

**Figure 4:**
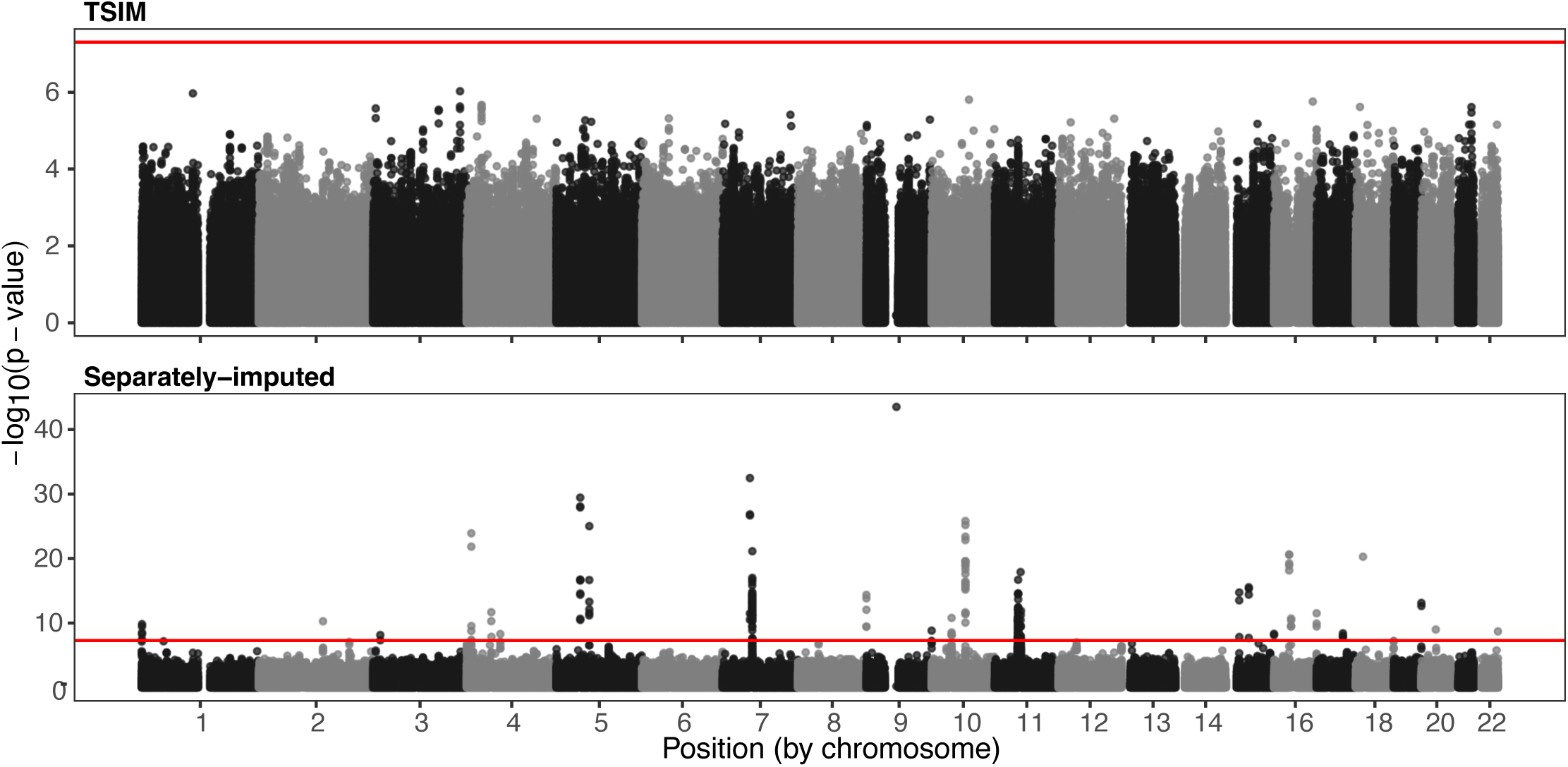
Manhattan plots for ART vs. UKB GWAS using cohort as outcome. Manhattan plots show GWAS results of the arthritis cohort (ART) controls vs. UKB samples. A simple merging method after separate imputations (*separately-imputed*, left) is compared to the TSIM method (*TSIM*). The *separately-imputed* results are filtered for R^2^ ≥ 0.80; the TSIM results are filtered for R^2^ ≥ 0.30. Red lines indicate the threshold for genome-wide significance (5×10^-8^).

**Figure 5:**
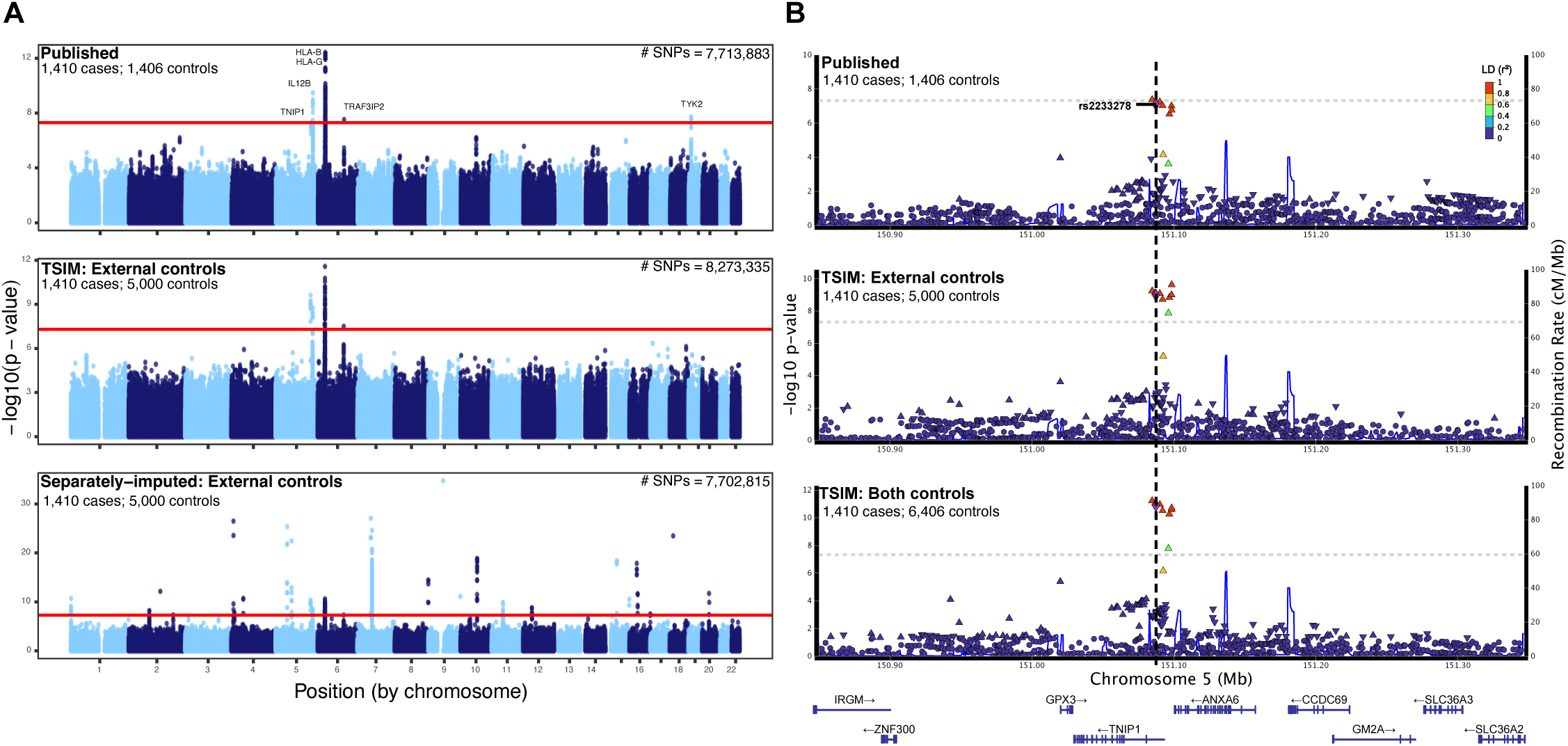
TSIM replicates psoriatic arthritis GWAS results and enables the use of case-only cohorts. (A) Manhattan plots show GWAS results of ART following common practices (*Published*: cases vs. internal controls, top), after merging cases from ART with UKB5K using TSIM (*TSIM:* cases vs. external controls, middle), and after single, separate imputations (*separately-imputed:* cases vs. external controls, bottom). Total number of SNPs are shown in upper right. Numbers of cases and controls are in the upper left of each plot. The *separately-imputed* results are filtered for R^2^ ≥ 0.80; the *Published* and *TSIM* results are filtered for R^2^ ≥ 0.30. Red lines indicate the genome-wide significance threshold (5×10^-8^). Genome-wide significant loci are labelled by nearest gene. See **Supp Fig 6** for Manhattan plots after merging cases and controls from arthritis cohort with UKB5K (cases vs. both controls). (B) Locus plots for *TNIP1* showing *TSIM* GWAS of cases vs. external controls (middle) and cases vs. both controls (bottom). GWAS results of cases vs. internal controls (top) is also shown.

### TSIM facilitates standard GWAS of multi-platform cohorts at the genotype level

Next, we evaluated whether a single joint GWAS of two European pSSNS cohorts using TSIM could accurately replicate results from a meta-analysis on the same data conducted by Barry et al.^17^. In the published analysis, these two cohorts (designated “EU” and “US” based on the geographical origin of each dataset) were meta-analyzed because they were genotyped on two distinct array platforms across five different sub-cohorts (**Supplemental Table S11**). When merged on hq-SNPs after the first stage of imputation, we found that the two cohorts clustered together in the PCA space (**Supplemental Fig. S8**), indicating that the samples had similar genetic population structure and might benefit from being analyzed together in a single TSIM joint GWAS (*TSIM Joint*). We also ran a TSIM meta-analysis (*TSIM Meta*), using TSIM to merge the four sub-studies comprising the EU cohort, to assess the impact of using TSIM to merge smaller cohorts. In total, there were 674 cases (EU=313, US=361) and 6,817 controls (EU=2,508, US=4,309). The TSIM GWAS included more SNPs (Joint=8,531,980, Meta=8,209,112) compared to the 8,014,298 included in the published GWAS and had similar results (**Fig. 6A**; **Supplemental Fig. S9**; **Supplemental Table S12,S13**). In both *TSIM Joint* and *TSIM Meta*, we replicated three out of four GWAS loci from the published meta-analysis (*HLA-DQB1* – rs17211699; *NFKBIL1* – rs2857607; *CALHM6* – rs2637678) (**Supplemental Table S13**). Using TSIM, we identified eight additional SNPs in high LD with rs2637678 at the *CALHM6* locus (**Fig. 6B**), expanding upon the 32 SNPs reported in the published analysis. Increasing coverage by including more SNPs in the GWAS improves the likelihood of capturing the true causal SNPs. The fourth published GWAS loci (*MORF4L1* – rs12911841) failed to achieve genome-wide significance after correcting for genomic control (**Supplemental Table S13**). This SNP had low imputation quality (R^2^ < 0.6) and the effect allele (T) frequency is 0.007 in Europeans^27^. Considering that the SNP is rare and has low imputation quality in our study population, we cannot conclude if TSIM shows an improvement by removing a false positive or if we simply lack the power to detect this association with this dataset. Nevertheless, TSIM provides a robust analysis of this multi-cohort pSSNS study. Furthermore, the low inflation and high concordance of *TSIM Joint* and *TSIM Meta* results compared to the published GWAS demonstrate that TSIM is an effective strategy for combining multiple cohorts and may be used as an alternative to meta-analysis for smaller cohorts, provided cohorts have similar genetic population structure.

**Figure 6:**
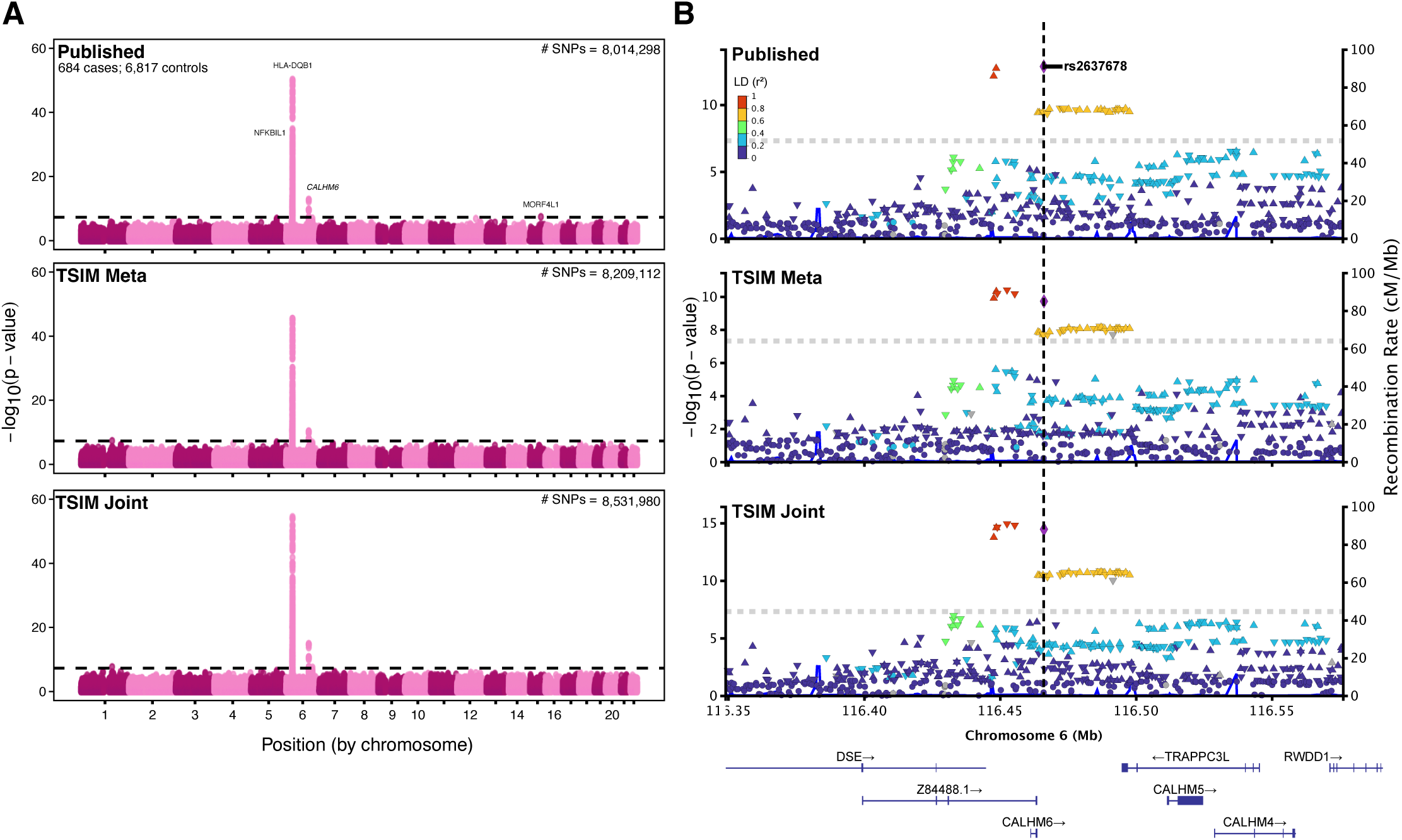
TSIM facilitates standard GWAS of multi-platform cohorts at the genotype level. (A) Manhattan plots show results of pSSNS European GWAS after merging data from five genotyping platforms using TSIM (*TSIM Joint*, bottom), meta-analysis of EU and US cohorts using TSIM to merge EU studies (*TSIM Meta*, middle), and the published meta-analysis (*Published*, top). Total number of SNPs are shown in upper right. 684 cases and 6,817 controls were used for all GWAS. Black dashed lines indicate the genome-wide significance threshold (5×10^-8^). Genome-wide significant loci are labelled by nearest gene. (B) Locus plot for *CALHM6* showing results of *TSIM Joint*, *TSIM Meta*, and *Published* GWAS.

## Discussion

We present TSIM, a method to extend the applicability of GWAS to previously understudied or underpowered cohorts. TSIM consists of two primary steps for efficiently harmonizing separately imputed cohorts bookended by two stages of imputation: (1) identify hq-SNPs and (2) merge cohorts based on hq-SNPs present in all cohorts. The identification of hq-SNPs corrects cohort-specific bias and the second stage of imputation increases the number of SNPs available for GWAS and downstream analyses. The development and evaluation of TSIM was done with data from UKB, a psoriatic arthritis case-control GWAS, and a pSSNS case-control GWAS meta-analysis. We evaluated imputation-derived error and GWAS results when implementing TSIM for two cohorts in our psoriatic arthritis analysis (ART and UKB) and for multiple cohorts in our pSSNS analysis (four in TSIM EU, five in TSIM Joint). In our validation, we showed that TSIM is an effective method to harmonize heterogeneous genotyping data with minimal increases in imputation-derived error. However, it cannot account for sample heterogeneity arising from differences in genetic population structure and other relevant phenotypic characteristics such as sex, age, and potential covariates for the phenotype of interest^1,3^. Similarly, in order to use TSIM for continuous phenotypes, robust harmonization of phenotype measurements must be performed separately.

TSIM addresses many issues with current practices for harmonizing genotype data which are insufficient and impractical for many datasets. In current practices, many SNPs are often removed when merging on the intersection and many samples may be dropped due to lack of matched controls. By merging separately imputed cohorts, TSIM enables the aggregation of case-only cohorts with external controls. TSIM also shows potential for combining datasets, at the genotype level, from different study centers to conduct a single joint GWAS analysis which performs just as well as a meta-analysis, as demonstrated with our pSSNS analysis. With the growing availability of public databases and biobanks, such as dbGaP^28^, UKB^29^, All of US^30^, FinnGen^31^, and others, TSIM offers the opportunity for researchers to apply GWAS to existing case-only cohorts using external controls. With the growing access and coverage of large biobanks and publicly available cohorts, our method provides an avenue through which cohorts for previously understudied phenotypes may be investigated in the GWAS framework.

TSIM also has some limitations, primarily the computational costs of imputing large genetic datasets twice and all the known limitations of imputation itself^9^. All cohorts should be imputed using the same reference panel and imputation algorithm. This may require researchers to run imputation on a large amount of data to ensure that all cohorts are processed appropriately. However, some biobanks, such as UKB, are imputing their data using publicly available reference panels and imputation methods, such as TOPMed, which may alleviate this burden for researchers in the future^10^. Additionally, because TSIM is largely reliant on high-quality imputation, the accuracy of the first stage of imputation and effectiveness of merging are dependent on key factors impacting imputation results, such as demographics and size of the reference panel and how well ancestry is matched to the study cohort. Thus, the optimal thresholds of R^2^, ER^2^, and HWE for defining hq-SNPs may differ depending on these factors. Ideally, there should be at least 300,000 hq-SNPs for the second stage of imputation^6^. If there are fewer hq-SNPs, the second stage of imputation may result in lower imputation quality and potentially more false positives in downstream analyses. We found that the TOPMed reference panel performed better for the African and European populations than East and South Asian. The size of the study sample may also impact imputation as smaller sample sizes are more likely to have less accurate imputation quality calculations resulting in inaccurate classification of hq-SNPs^9,32,33^. Many of these issues with imputation have been addressed in previous research. For instance, Sun et al. have developed MagicalRsq^32^, a machine-learning-based genotype imputation quality calibration, which takes a sample size agnostic approach to calculating imputation quality. Additionally, meta-imputation, which combines imputation results from multiple imputations with different reference panels, has been shown to improve imputation accuracy and results, especially for admixed individuals^34^. Both these methods may be implemented in TSIM after or during the first stage of imputation, respectively, and before identifying hq-SNPs.

There are two data processing steps which will have major impacts on results. First is the pre-imputation QC. With two stages of imputation, there is the potential for genotype errors made in the raw data to propagate if insufficient QC is done. We have found that running “Imputation preparation and checking” from the McCarthy Group Tools (https://www.chg.ox.ac.uk/~wrayner/tools/) as well as filtering out poorly genotyped SNPs is crucial to ensure homogeneity among all cohorts-to-be-merged. Second is to keep in mind any pre-processing that was done on the data, particularly when the cohorts are received in different formats. For example: PLINK^35^, a tool often used for QC in GWAS, codes minor and major alleles based on allele frequency while VCF files code reference and alternative alleles based on a reference. As the reference allele is not always the major allele, some SNPs could have flipped allele codes when attempting to merge cohorts. Fortunately, these biases may also be mitigated by McCarthy Group Tools.

Additionally, we only evaluated TSIM GWAS results in psoriatic arthritis and steroid sensitive nephrotic syndrome, both diseases with binary classification that have a small number of large effect loci that can be detected with relatively small samples. Additional research applying TSIM to other datasets involving different genotyping platforms, genetic population structures, and phenotypes will be beneficial to further understand TSIM’s limitations and applications.

## Methods

### Two-stage imputation method (TSIM)

In TSIM, we first impute each cohort separately using the same reference panel and imputation algorithm, and then high-quality genotyped and imputed SNPs (hq-SNPs) are identified. Then, the cohorts are merged based on SNPs present in all hq-SNPs sets. Lastly, this merged dataset undergoes a second stage of imputation, after which post-imputation analysis proceeds as normal. With the TOPMed imputation server, hq-SNPs are defined as SNPs with imputation quality (R^2^) ≥ 0.98, minor allele frequency (MAF) ≥ 0.01, Hardy-Weinberg equilibrium (HWE) p-value ≥ 1×10^-6^ for controls, and, if genotyped, empirical imputation quality (ER^2^) ≥ 0.9. Our command line tool, tsim, implements these two steps (i.e., hq-SNPs identification and merging) and outputs per chromosome VCFs ready for imputation. Additional sample QC to remove outliers or unmatched cases or controls may be done on merged unimputed genotypes if there is sufficient SNP overlap or following cohort merging on hq-SNPs after the first stage of imputation. The TOPMed Imputation Server v2.0.0-beta3 (Minimac4 for imputation, Eagle v2.4 for phasing, r3 for reference panel) was used for all our analyses, with the exception of the pSSNS GWAS which was run using TOPMed Imputation Server v1.6.6 (Minimac4, Eagle v2.4, and r2 reference panel).

### Quality control

Before imputation, all cohorts underwent similar quality control using PLINK 1.9^35^. SNPs with missingness ≥ 0.02, MAF < 0.01, and/or HWE p-value ≤ 1×10^-6^ for controls were removed. Samples with heterozygosity greater than four standard deviations from mean, missingness ≥ 0.02, closer than second-degree relation to other samples, and/or outlying in the principal component analysis (PCA) space (by visual inspection) were also removed. Following imputation, we filtered out low-quality and rare imputed variants from all datasets. This included variants with R^2^ < 0.3, MAF < 0.01, HWE p-value ≤ 1×10^-6^ for controls, and if genotyped, ER^2^ < 0.9. Further quality control was done according to the analysis conducted (see following methods for specific details on each analysis).

### Determination of high-quality SNPs

We evaluated the imputation-derived error for SNPs with high imputation quality (R^2^ ≥ 0.95) stratified by R^2^ bins. Only SNPs with R^2^ ≥ 0.95 and MAF ≥ 0.01 in the UK Biobank (UKB) imputed datasets were analyzed and each ancestry group (defined by UKB’s genomic ancestry labels: European (EUR), African (AFR), Admixed American (AMR), East Asian (EAS), and South Asian (SAS)) was analyzed separately. Here, we conducted two different analyses, one using WES (200k release) and the other using WGS (ML-corrected DRAGEN population level WGS variants, pVCF format, 500k release) genotypes to represent “ground truth.” Each of the five ancestry groups were evaluated separately. For AFR, AMR, EAS, and SAS, all available samples that passed sample QC metrics (described above) and had WES sequencing were used for analysis. For EUR, we first combined all UKB batches and performed sample and SNP QC on the merged dataset. Then, we subset EUR samples to UKB batches 5, 35, 65, and 95, consisting of 15,495 samples. Among these, we further selected samples for which WES data are available (n=6,911) for hq-SNPs and imputation-derived error analysis and a random subset of 5,000 for our GWAS analyses. For analyses utilizing WGS, samples lacking WGS data were removed. For each ancestry group, we filtered WES/WGS for good quality biallelic SNPs with MAF ≥ 0.01, PASS in the FILTER column of the VCF, and sample missingness < 0.01. The imputed dosages were compared to their WES and WGS genotypes using GLIMPSE2, which computes overall genotyping error rate^21,22^. We ran GLIMPSE2 with the following binning options: `--bins 0.005 0.010 0.020 0.050 0.100 0.200 0.500`. In addition, for WES comparison, we added the following options to remove lower quality genotypes: `--min-val-dp 8 --min-val-gl 0.9999`.

### Evaluating imputation-derived error

Unrelated samples from UKB with array-based genotyping on UKB Axiom, WES, and WGS were used to evaluate imputation-derived error. Similar to the analysis of high-quality SNPs as described above, we used GLIMPSE2 with the same parameters for genotype comparison. We use S1 to refer to results from the first imputation and S2 to refer to results from the second imputation. We stratified per SNP calculations into R^2^ bins based on the imputation quality. This analysis was restricted to variants that were present in either S1 or S2 imputations with MAF ≥ 0.01, R^2^ ≥ 0.30, ER^2^ ≥ 0.90, and in our “good quality” WES/WGS SNP set.

### Investigating impact of TSIM on type I Error

We used a subset of 5,000 EUR samples from UK Biobank (UKB) and 1,406 controls from ART (see below) to conduct a control vs. control GWAS. ART and UKB samples were projected to the 1KGP PCA space using KING^36^ to harmonize ancestry group annotations. All datasets underwent the standard quality control process as described above (under *Quality control*). SAIGE^37^ v0.29.5 was used to conduct GWAS analysis on dosages and to account for case-control imbalances and 5 principle components were included as covariates. For SAIGE Step 1, - -LOCO=FALSE was used. For Step 2, the following parameters were used: --minMAF=0.01, --minMAC=1, --LOCO=FALSE. LocusZoom^38^ was used for visualization.

### Investigating impact of TSIM on published psoriatic arthritis GWAS results

The psoriatic arthritis study cohort (ART), containing both cases and controls, came from a published GWAS meta-analysis on psoriatic arthritis genotyped on the HumanOmni1-Quad BeadChip^16^. We obtained the genotype data through dbGaP controlled data access. All study cohort samples were annotated as Caucasian. We randomly selected 5,000 individuals from 427,234 Europeans in UKB. Individuals with psoriatic arthritis or failing QC were removed prior to random selection. This subset of UKB individuals comprised our external control cohort genotyped on UKB Axiom (UKB5K). There were 1,410 cases and 6,406 controls (1,406 from the study cohort; 5,000 from UKB5K) passing QC.

We conducted two separate analyses, the ART vs. UKB control GWAS (see above) and evaluating TSIM using GWAS results from two different scenarios using different combinations of our cohorts:

1. *both controls*: ART cases and controls merged with UKB5K
2. *external controls*: ART cases merged with UKB5K

For *separately-imputed* GWAS, cohorts were merged after single, separate imputations (Stage 1). Following the first stage of imputation, we first filtered for SNPs with R^2^ ≥ 0.30 and MAF ≥ 0.01, then merged the cohorts on intersecting SNPs. For *TSIM* GWAS, cohorts were merged using TSIM. We also conducted a GWAS of only ART cases and controls to generate reference results for comparison (*internal-controls*). All datasets underwent the standard quality control process as described above. SAIGE^37^ v0.29.5 was used to conduct GWAS analysis on dosages and account for case-control imbalances, using the parameters described above. All reported p-values were adjusted for genomic control to accurately compare results. LocusZoom^38^ was used to create locus plots in **Supp Fig 7.**

Of note, it was not feasible to perform a GWAS using a common practice, which combines separate cohorts before imputation, since ART and UKB were genotyped separately on two different genotyping platforms intersecting by only 100,000 SNPs after QC. This is insufficient coverage for accurate genotype imputation^6^.

### Replicating results from pSSNS GWAS meta-analysis

We applied TSIM and replicated results from a pediatric steroid-sensitive nephrotic syndrome (pSSNS) GWAS meta-analysis our lab previously published^17^. Specific details on the published pSSNS GWAS methods and cohort composition can be found in Barry et al.^17^. Briefly, our pSSNS GWAS consisted of a meta-analysis of two European cohorts consisting of five separate genotype batches with different collection locations. Samples were projected to 1KGP PCA space to infer ancestry using PEDDY^39^. The European Union (EU) cohort consists of four sub-cohorts: case and control data from Sorbonne Université in Paris and the NEPHROVIR study^40^, as well as healthy controls from the Three Cités Study^41^ and 1KGP. The United States (US) cohort contained cases and controls obtained from Columbia University in New York^17^. For EU, all individual studies were imputed separately in the first stage and then combined before the second stage of imputation with the US cohort. See **Supp Table 3** for more details on each study.

We evaluated TSIM in two different analyses: (1) GWAS was run separately in EU (using TSIM to merge studies) and US cohorts, then combined in a meta-analysis using the STDERR scheme in METAL^42^, and (2) all studies in both EU and US cohorts were merged using TSIM and analyzed in one joint GWAS. For both analyses, PLINK 1.9^35^ was used to conduct GWAS on genotypes in order to better compare to the published results which also used PLINK. All reported p-values were adjusted for genomic control to better compare results. LocusZoom^38^ was used to create locus plots in **Supp Fig 9.**

## Data Availability

All data produced in the present study are available upon reasonable request to the authors.

https://github.com/dongwonlee-lab/tsim

## Data Availability

Data used included genotype data from UK Biobank, dbGaP study phs000982.v1.p1, and data used in our previous pSSNS GWAS.

## Code availability

The tsim command line tool is available at https://github.com/dongwonlee-lab/tsim.

## Acknowledgements

This research has been conducted using UK Biobank Resource under Application Number 64945. We would like to acknowledge the Boston Children’s Hospital High-Performance Computing Resources BCH HPC Clusters, Enkefalos 3 (E3), made available for conducting the research reported in this publication. Software used in the project was installed and configured by BioGrids^43^. We would like to also thank Pierre Ronco and Simone Sanna-Cherchi for allowing us to use the pSSNS dataset in our analysis.

