## Supplemental Information for "Accurate cross-platform GWAS analysis via two-stage imputation"

### A method to improve GWAS by combining cohorts across genotyping platforms

Includes:

- Supplemental figures S1-S9 with legends
- Legends for supplemental tables S1-S13

### Supplemental Figures

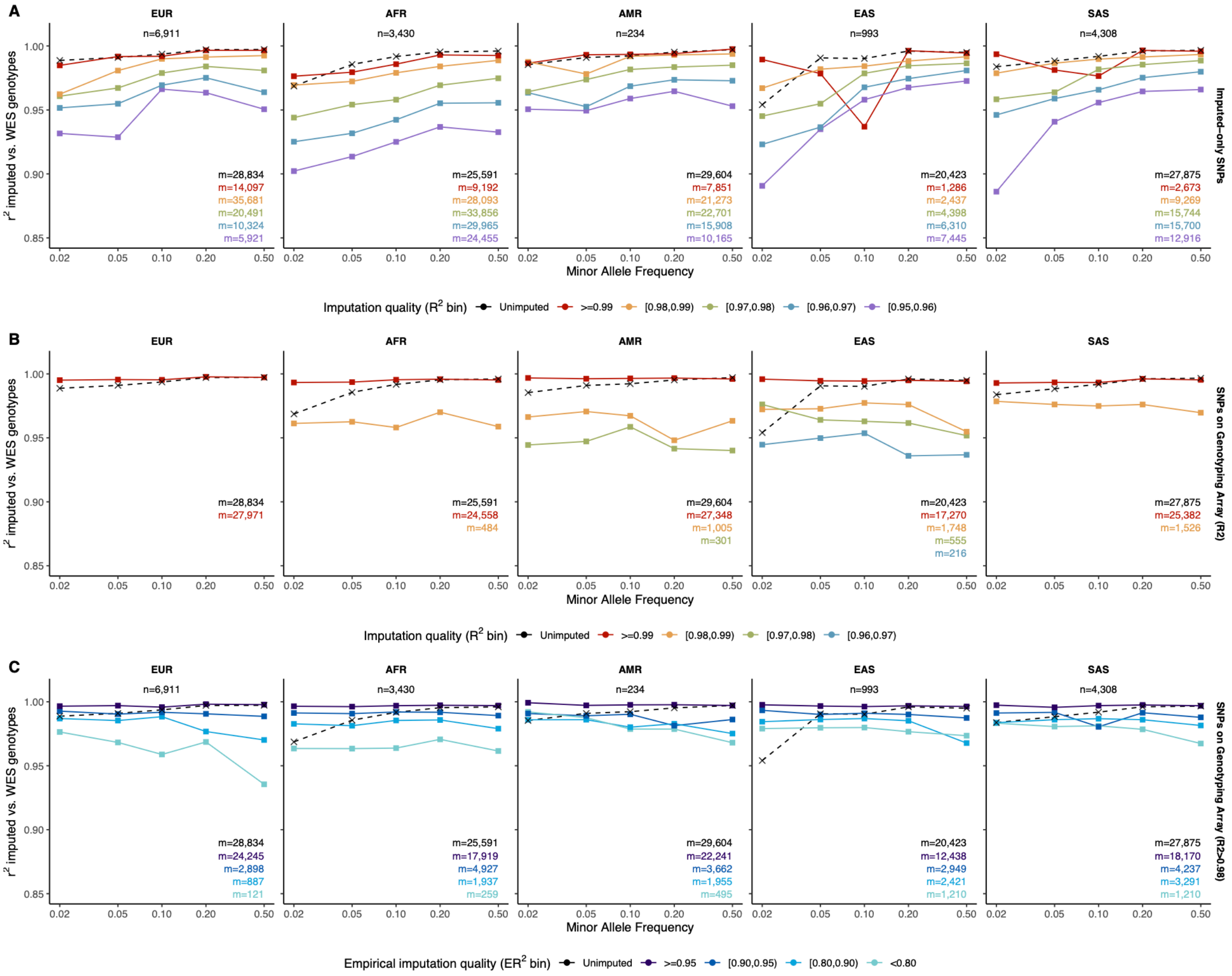


**Supplemental Figure S1: Determination of high-quality SNPs (hq-SNPs) using WES**
Similar to **Fig. 2**, line plots show correlation between imputed and WES genotypes as a function of MAF after the first stage of imputation stratified by whether SNPs were **(A)** imputed-only, **(B)** present in genotyping array, or **(C)** present in genotyping array with $R^2 \geq 0.98$ for UKB Europeans (EUR), Africans (AFR), Admixed Americans (AMR), East Asians (EAS), and South Asians (SAS). SNPs with $R^2 \geq 0.95$ were included, which were then stratified by imputation quality into $R^2$ bins (A-B) or empirical $R^2$ ($ER^2$) (C). Unimputed (with $ER^2 \geq 0.9$) vs. WES (dotted line) shows correlation between unimputed SNP genotypes compared to WES for comparison. The total number of samples is *n* (top center) and the total number of SNPs in each stratum is *m* and colored by $R^2$ bins (lower right). The y-axis shows the correlation and the x-axis shows the MAF.

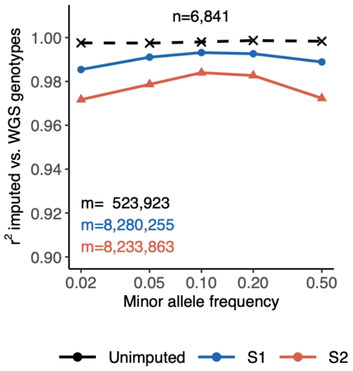


**Supplemental Figure S2: TSIM shows comparable imputation-derived error for S1 and S2**
Similar to **Fig. 3**, line plots show correlation between imputed and WGS genotyped as a function of MAF after the first (S1, blue) and second (S2, red) stages of imputation for UKB Europeans (EUR). SNPs with imputation quality ($R^2$) ≥ 0.30 were included. Unimputed (with empirical $R^2 \geq 0.9$) vs. WGS (dotted line) shows correlation between unimputed SNP genotypes compared to WGS for comparison. The total number of SNPs in each stratum is *m* and colored by stage (lower left). The y-axis shows the correlation and the x-axis shows the MAF.

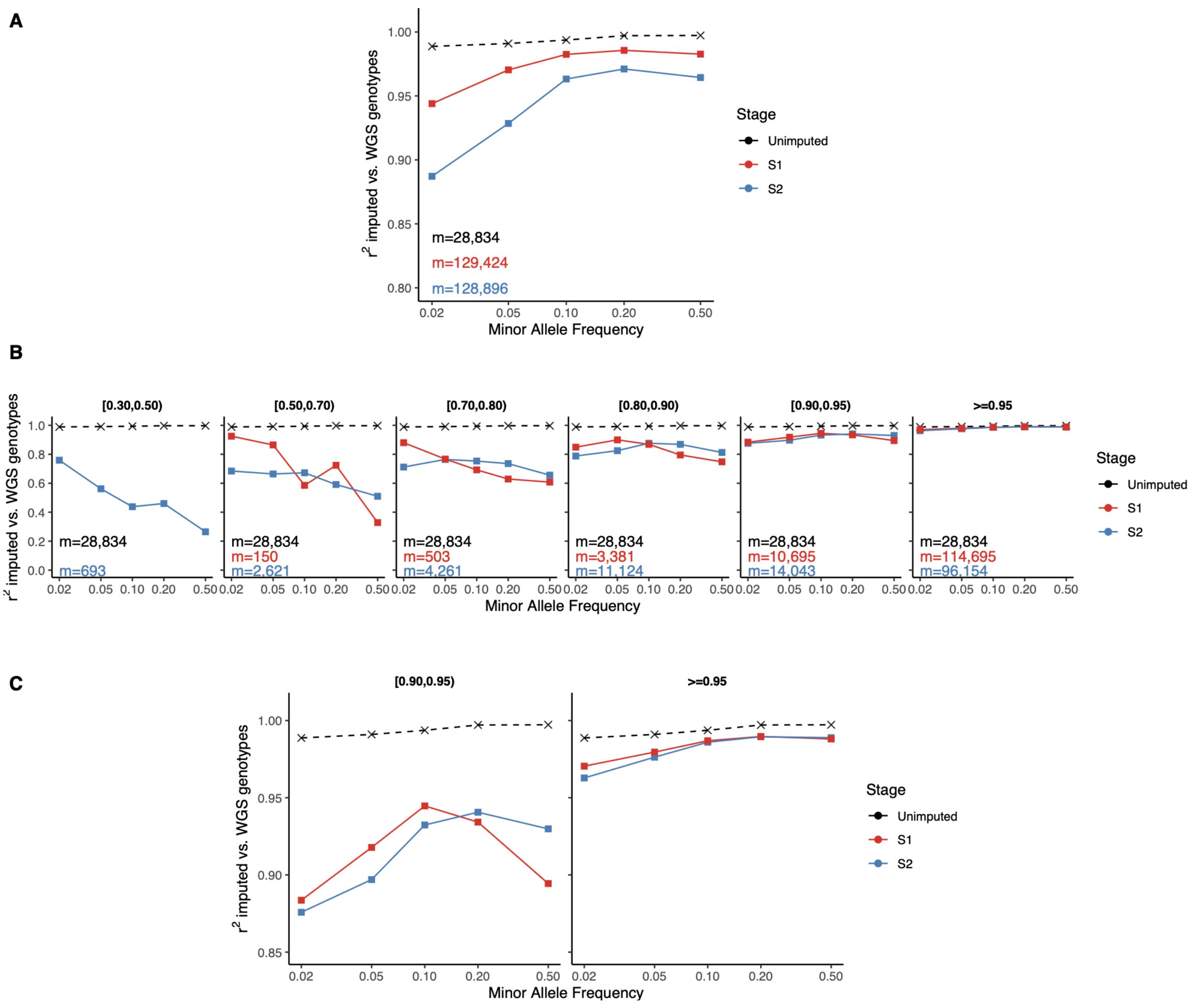


**Supplemental Figure S3: TSIM shows comparable imputation-derived error for S1 and S2 using WES** Similar to **Fig. 2** and **Supplemental Fig. S2**, line plots show correlation between imputed and WES genotyped as a function of MAF after the first (S1, red) and second (S2, blue) stages of imputation **(A)** overall and **(B,C)** stratified by imputation quality ($R^2$) for UKB Europeans (EUR). (B) Shows all $R^2$ bins while (C) shows a zoomed in version for $R^2$ bins of [0.90,0.95) and >=0.95. SNPs with $R^2 \geq 0.30$ were included, which were then stratified by imputation quality into $R^2$ bins. Unimputed (with empirical $R^2 \geq 0.9$) vs. WES (dotted line) shows correlation between unimputed SNP genotypes compared to WES for comparison. The total number of SNPs in each stratum is *m* and colored by $R^2$ bins (lower left). The y-axis shows the correlation and the x-axis shows the MAF.

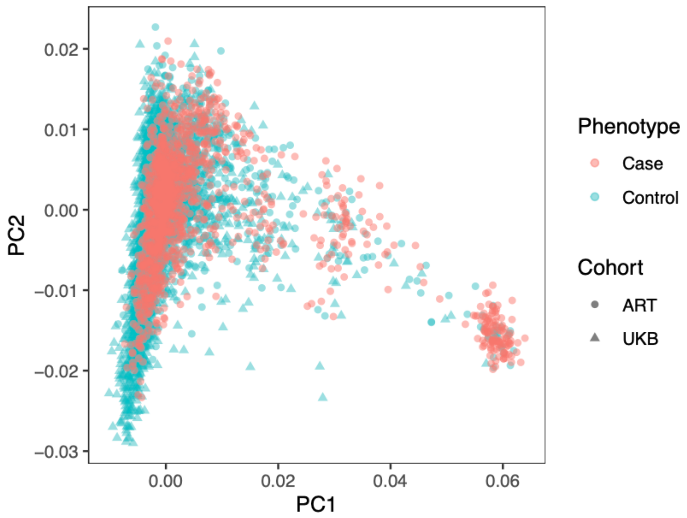


**Supplemental Figure S4: PCA for arthritis and UKB**
PCA plot for GWAS of arthritis and UKB5K.

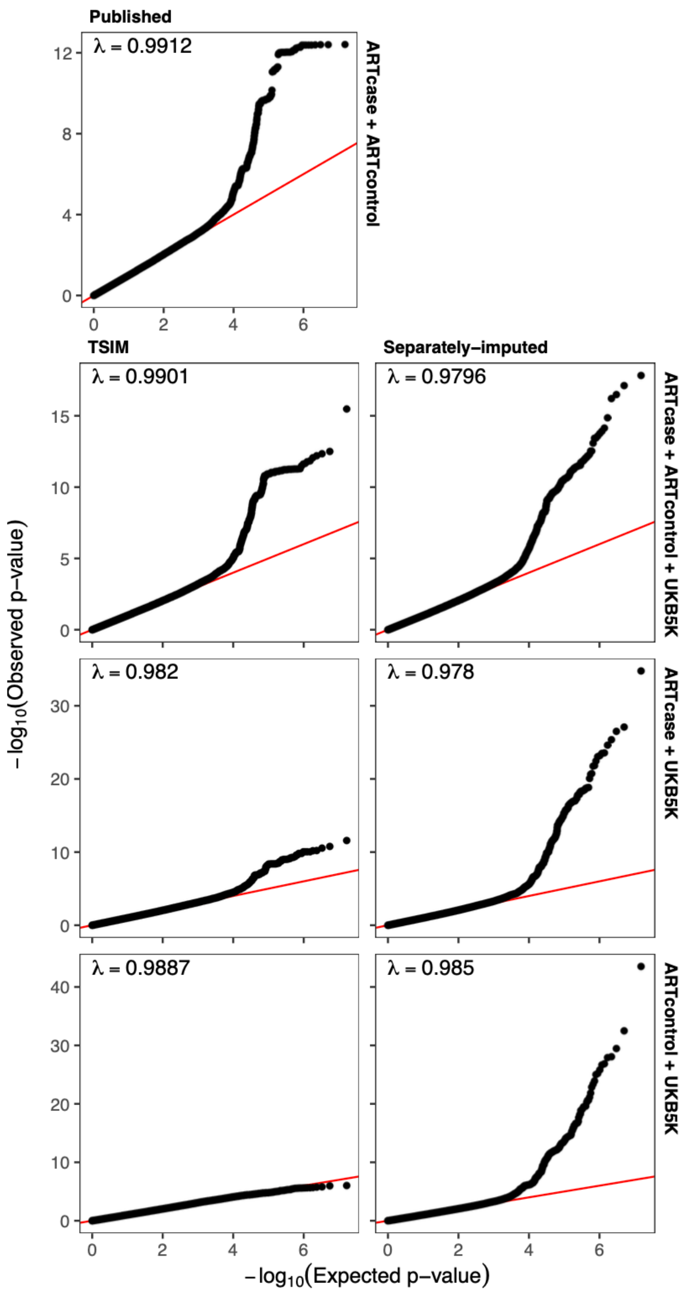


**Supplemental Figure S5: QQ plots for arthritis and UKB GWAS**

QQ plots show GWAS results of arthritis and UKB for three different scenarios. Scenarios include arthritis cases merged with UKB5K (2nd row), arthritis cases and controls merged with UKB5K (3rd row), and arthritis controls merged with UKB5K (bottom). *Published* GWAS results of only arthritis cohort shown in 1st row. For each scenario, TSIM (*TSIM*, right) is compared to a simple merging method after separate imputations (*separately-imputed*, left). Genomic inflation factor (λ) is shown in upper left.

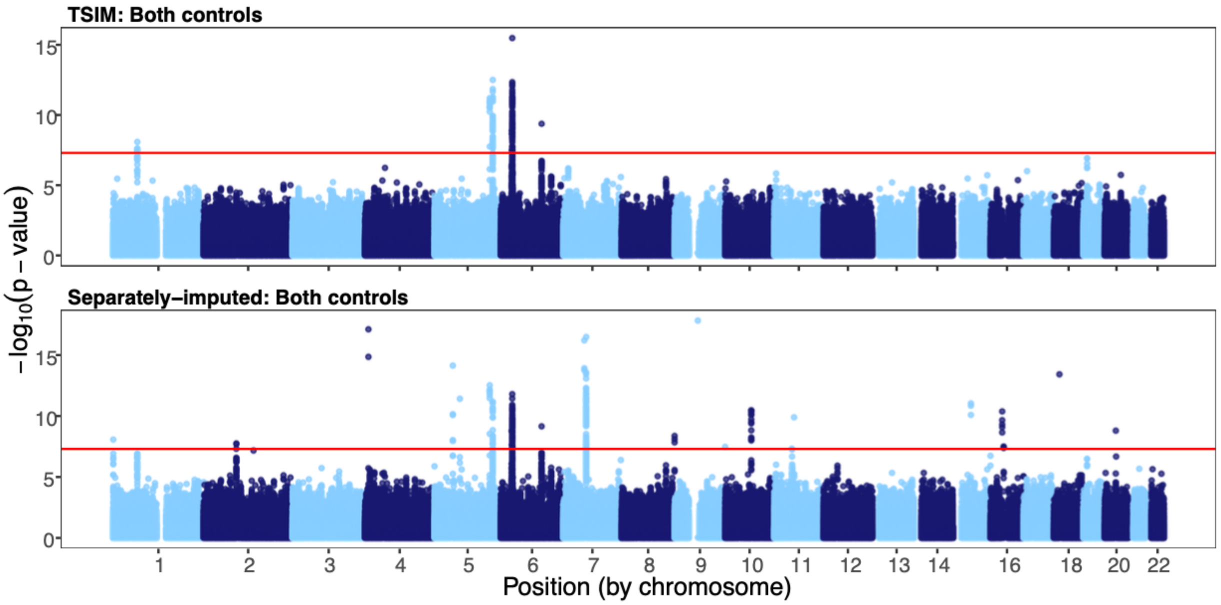


**Supplemental Figure S6: Manhattan plots for GWAS of arthritis cases and controls merged with UKB**
Manhattan plots show GWAS results of merging cases and controls from the arthritis cohort with UKB5K using TSIM (*TSIM*, top) and a simple merging of separately imputed data *(separately-imputed*, bottom). Red lines indicate the threshold for genome-wide significance ($5\times10^{-8}$).

**A *HLA-B***

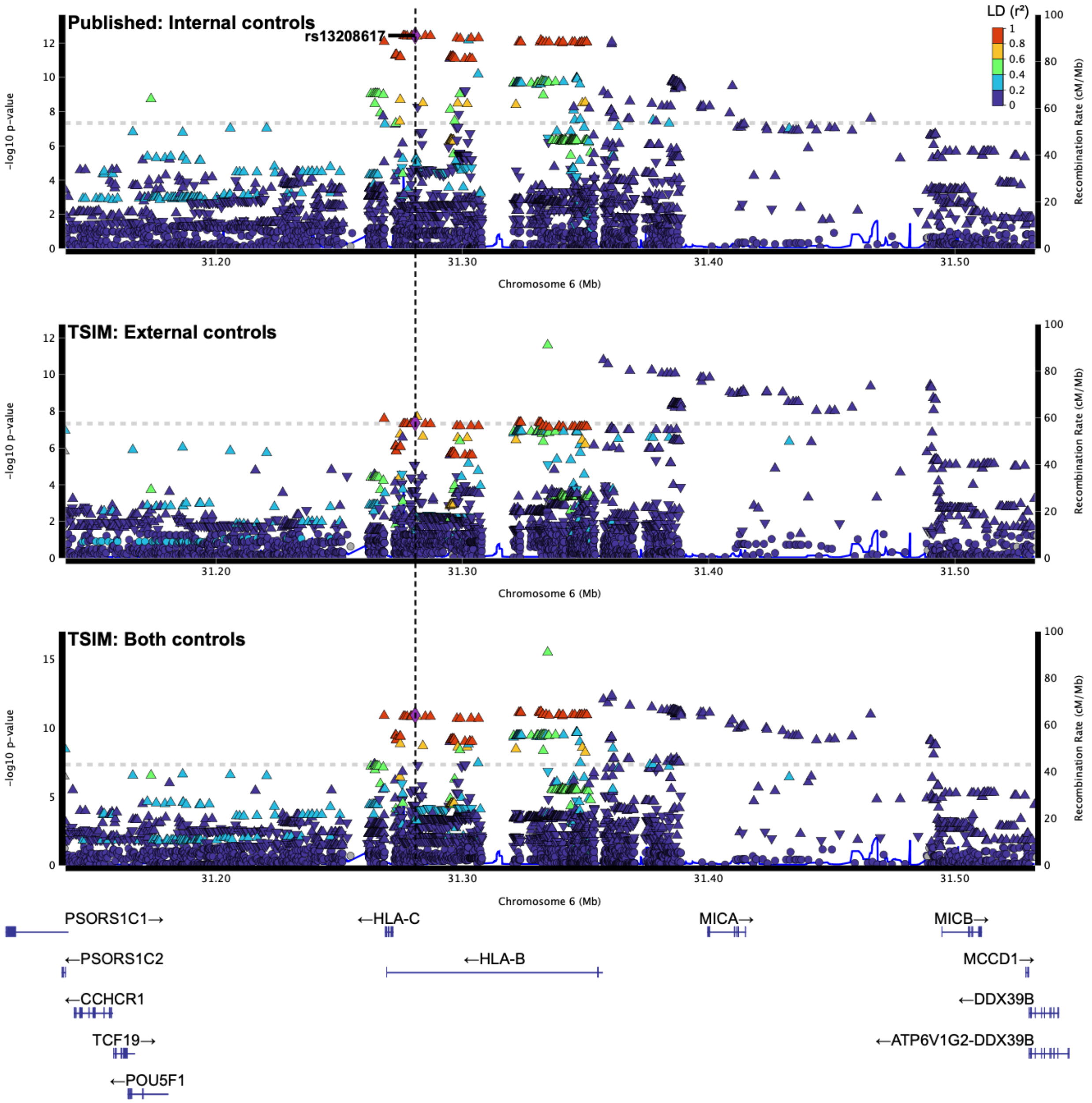
Published: Internal controls
rs13208617
LD (r²)
1
0.8
0.6
0.4
0.2
0
−log10 p-value
Recombination Rate (cM/Mb)
Chromosome 6 (Mb)
31.20
31.30
31.40
31.50
TSIM: External controls
TSIM: Both controls
PSORS1C1→
←PSORS1C2
←CCHCR1
TCF19→
←POU5F1
←HLA-C
←HLA-B
MICA→
MICB→
MCCD1→
←DDX39B
←ATP6V1G2-DDX39B

**B *IL12B***

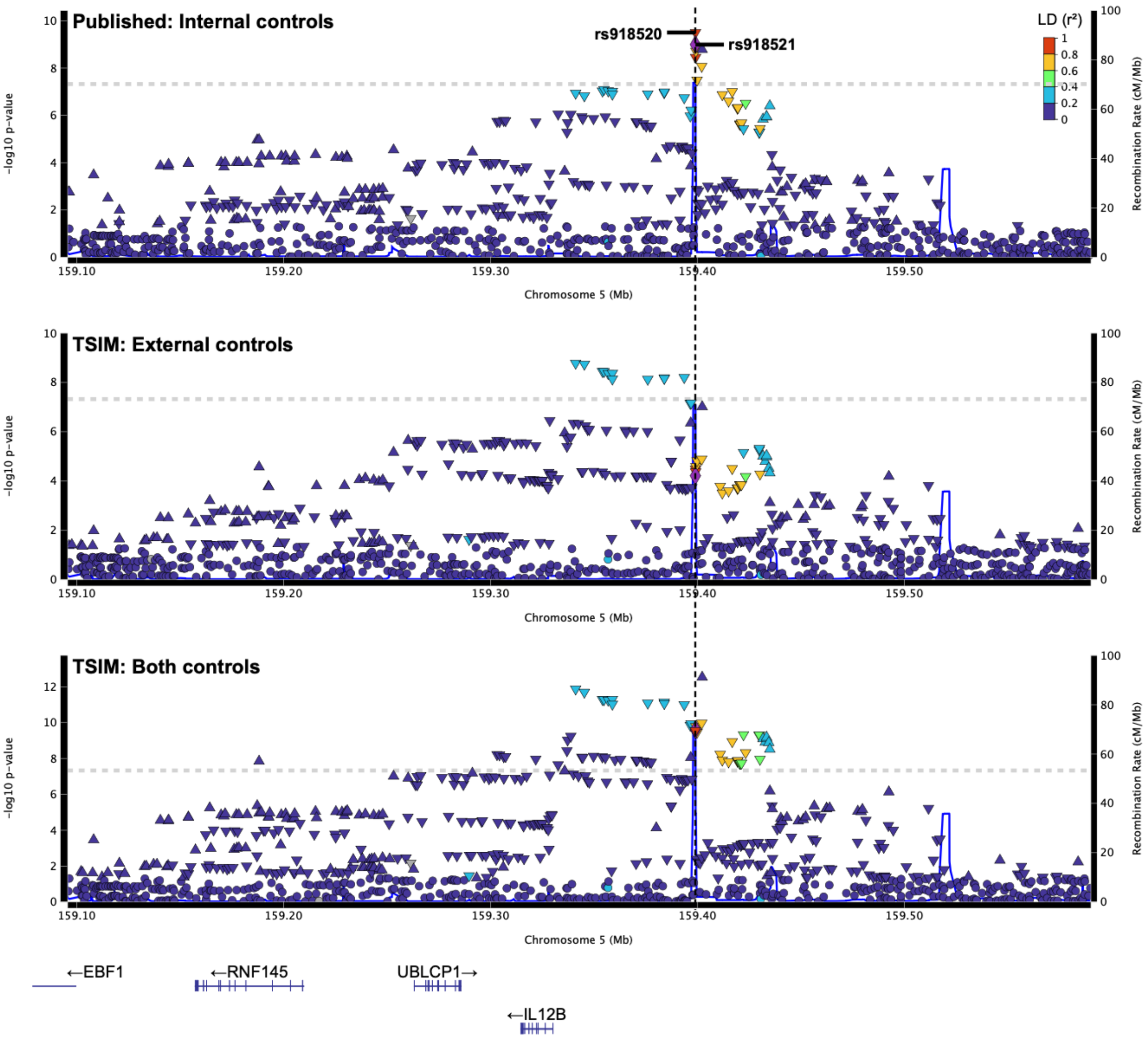

Published: Internal controls
rs918520
rs918521
LD (r²)
1
0.8
0.6
0.4
0.2
0
-log10 p-value
Recombination Rate (cM/Mb)
Chromosome 5 (Mb)
159.10
159.20
159.30
159.40
159.50
TSIM: External controls
TSIM: Both controls
←EBF1
←RNF145
UBLCP1→
←IL12B

**C *TYK2***

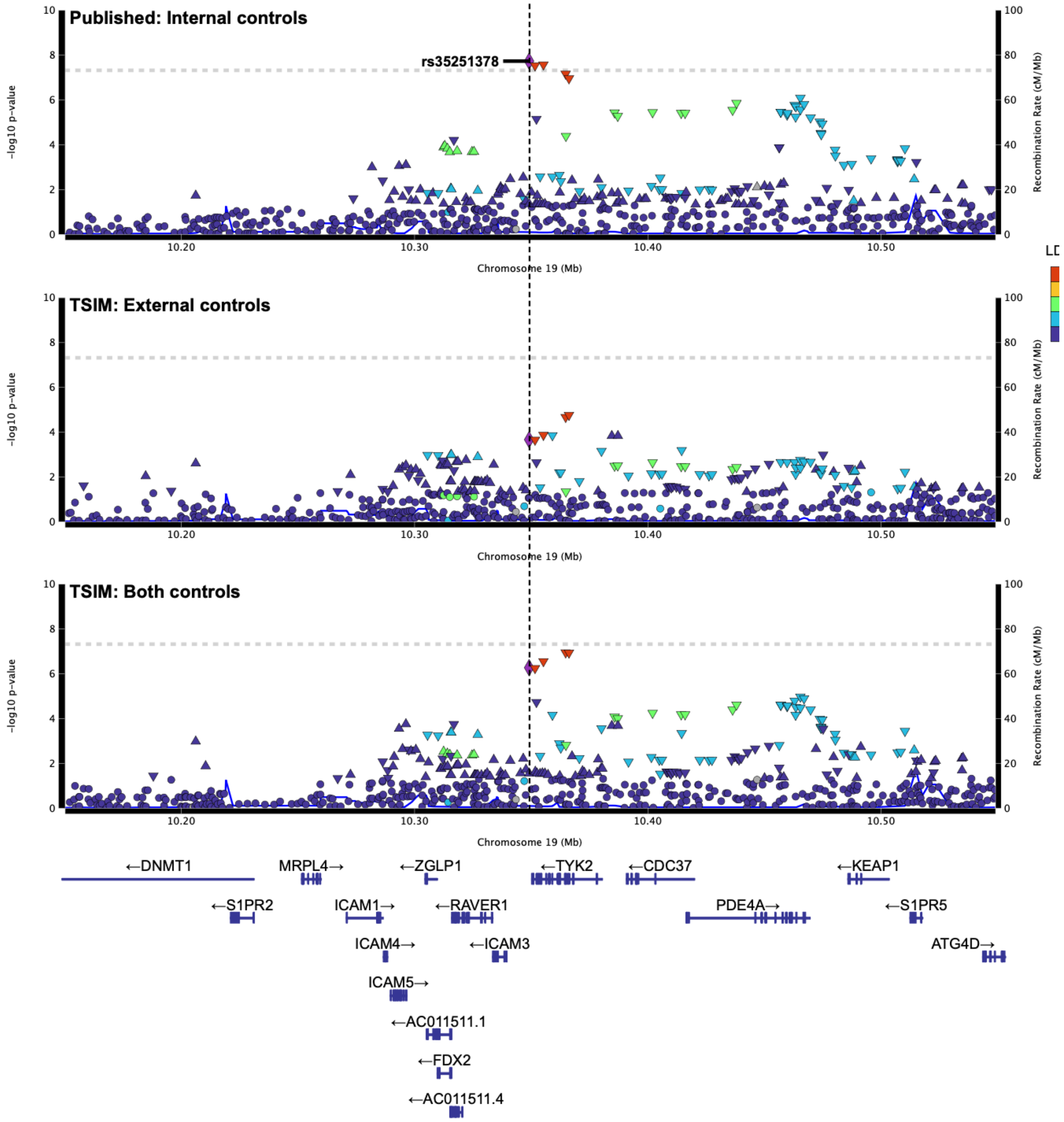

Published: Internal controls
rs35251378
TSIM: External controls
TSIM: Both controls
-log10 p-value
Recombination Rate (cM/Mb)
Chromosome 19 (Mb)
10.20
10.30
10.40
10.50
LD
←DNMT1
MRPL4→
←ZGLP1
←TYK2
←CDC37
←KEAP1
←S1PR2
ICAM1→
←RAVER1
PDE4A→
←S1PR5
ICAM4→
←ICAM3
ATG4D→
ICAM5→
←AC011511.1
←FDX2
←AC011511.4

#### D *TRAF3IP2*

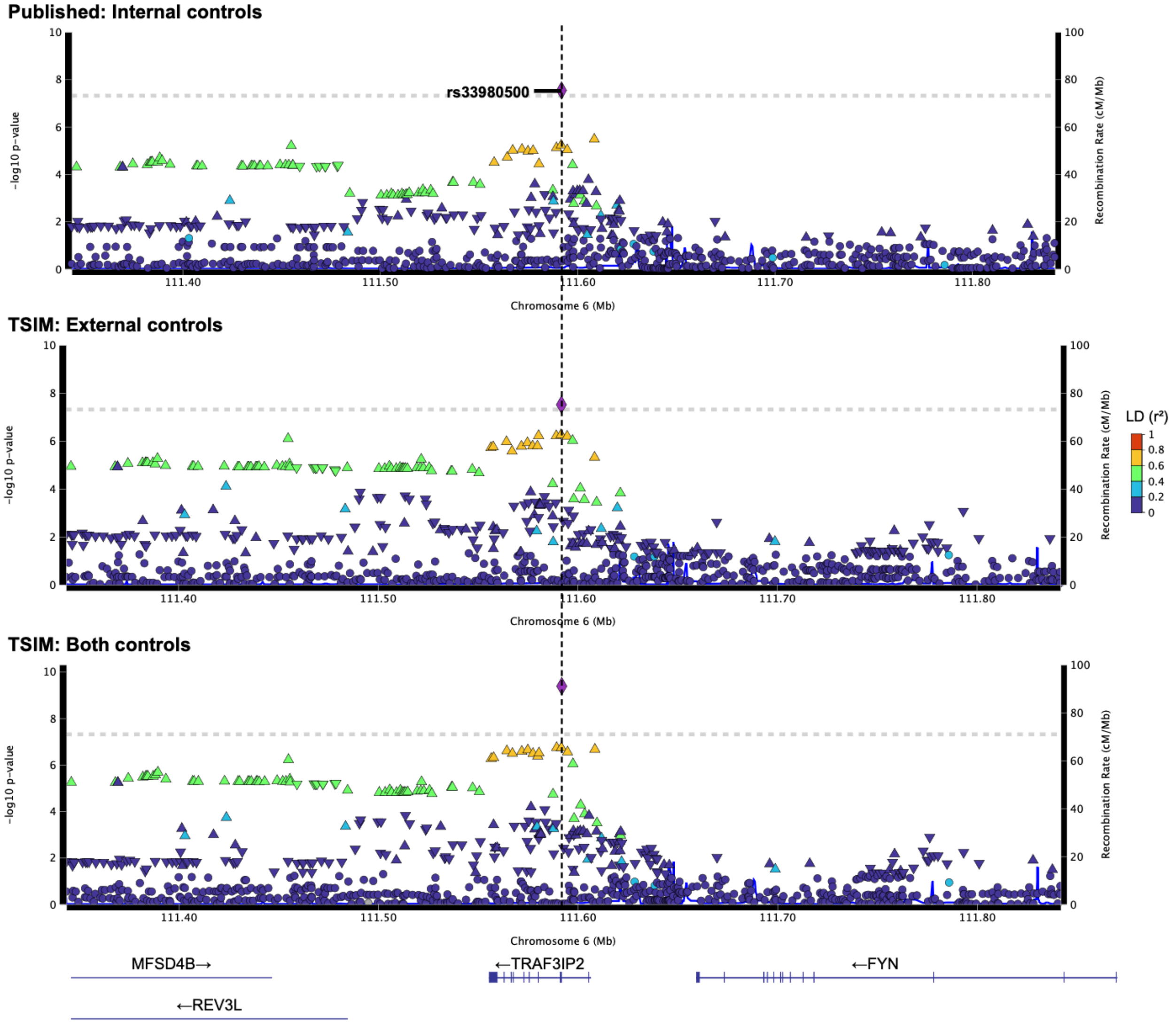
Published: Internal controls
rs33980500
TSIM: External controls
TSIM: Both controls
-log10 p-value
Recombination Rate (cM/Mb)
Chromosome 6 (Mb)
111.40
111.50
111.60
111.70
111.80
LD (r²)
1
0.8
0.6
0.4
0.2
0
MFSD4B→
←REV3L
←TRAF3IP2
←FYN

#### E *HLA-G*

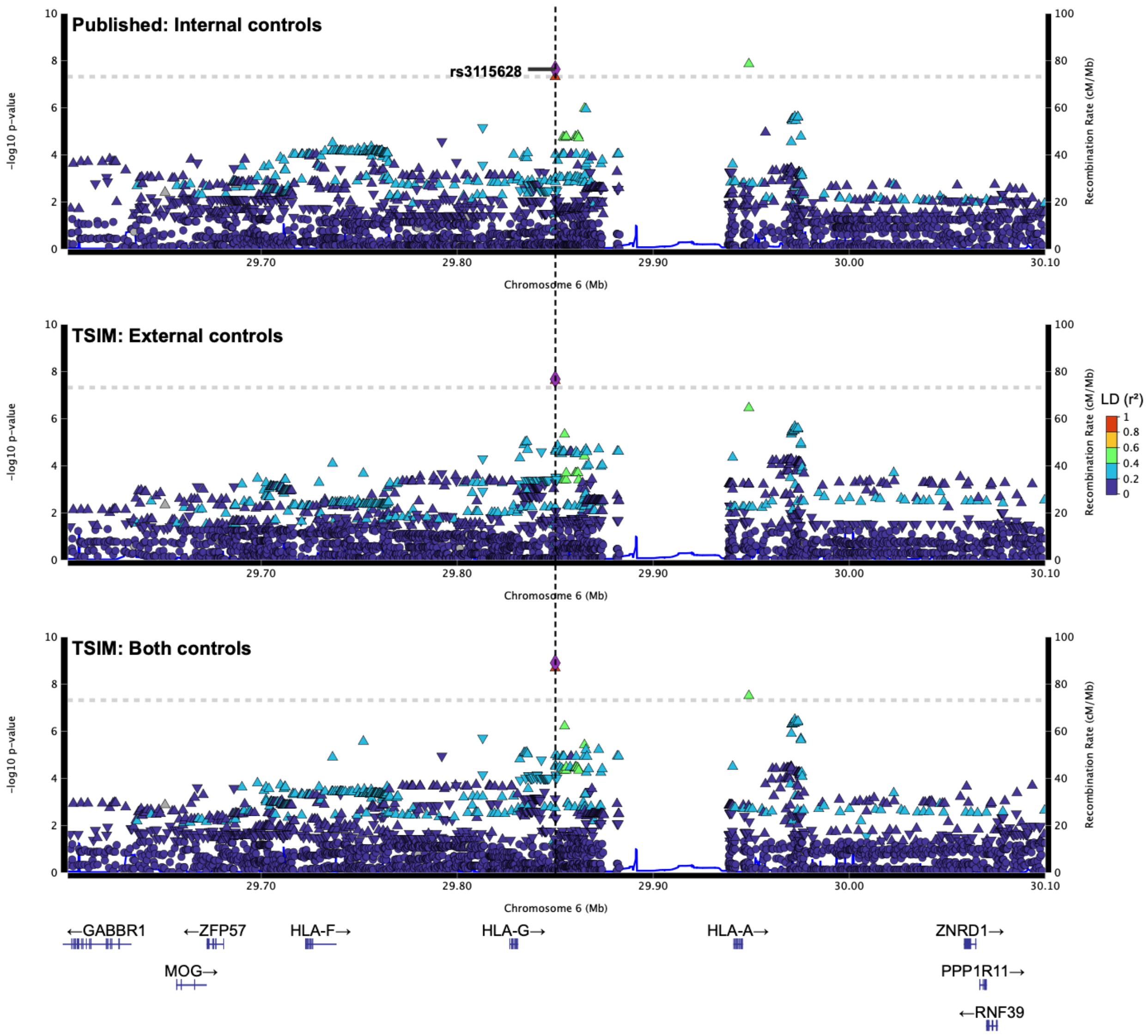


**Supplemental Figure S7: Locus plots for GWAS loci of Arthritis cohorts merged with UKB**
Locus plots highlighting SNPs detailed in **Supplemental Table S10** for *Published* GWAS (top), *TSIM external controls* (middle), and *TSIM both controls* (bottom) scenarios. Gene loci include **(A)** *HLA-B*, **(B)** *IL12B*, (C) *TYK2*, **(D)** *TRAF3IP2*, and **(E)** *HLA-G*.

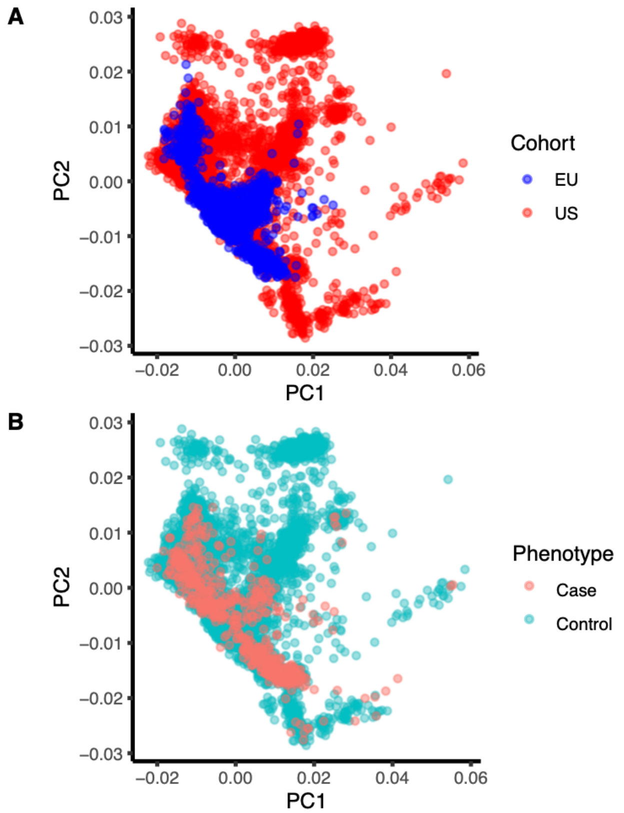


**Supplemental Figure S8: PCA of TSIM-merged EU and US cohorts from pSSNS GWAS**
PCA analysis of EU and US cohorts from pSSNS GWAS merged using TSIM colored by **(A)** cohort and **(B)** phenotype.

A *HLA-DQB1*

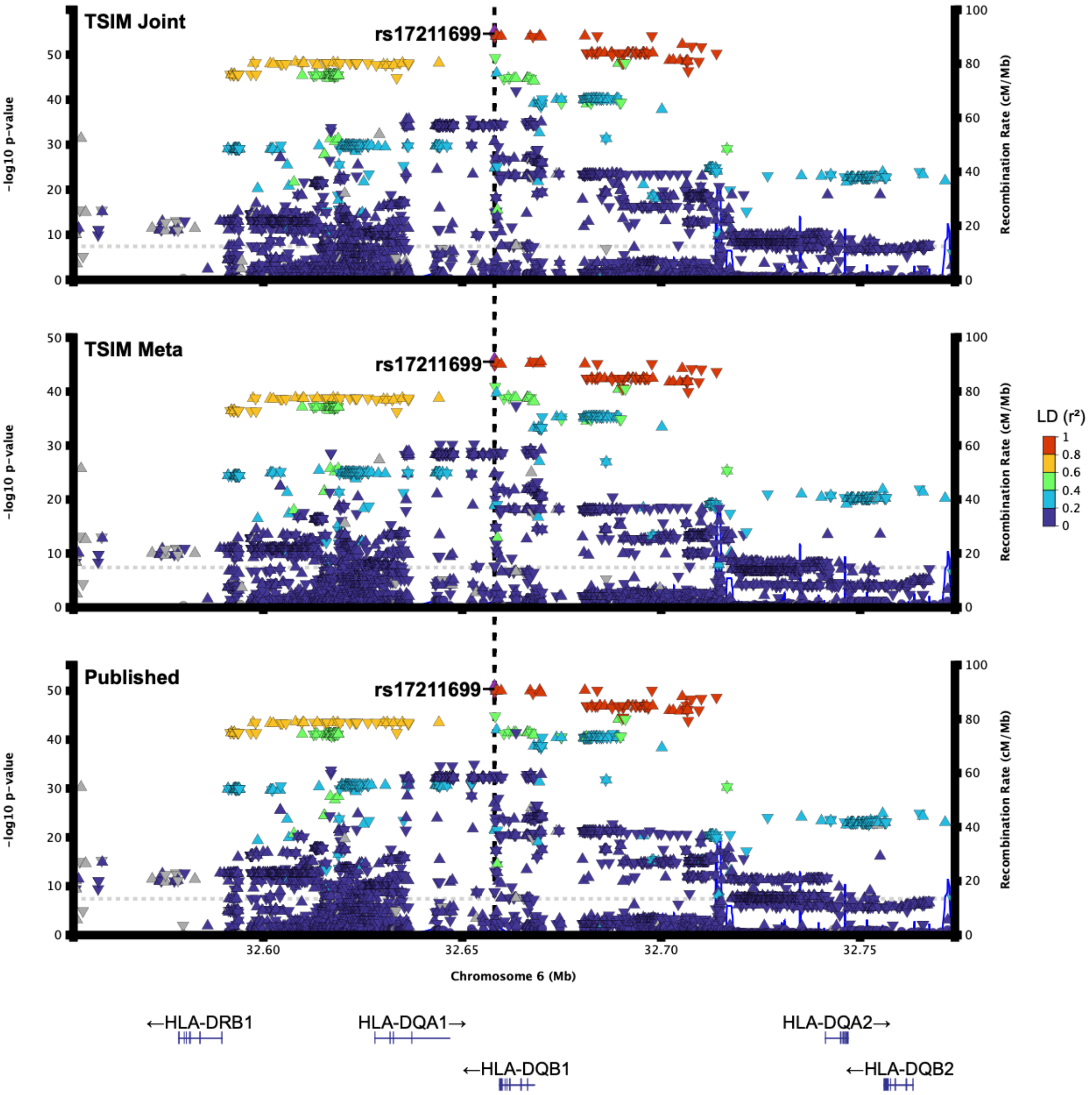

TSIM Joint
TSIM Meta
Published
rs17211699
-log10 p-value
Recombination Rate (cM/Mb)
LD (r²)
1
0.8
0.6
0.4
0.2
0
32.60
32.65
32.70
32.75
Chromosome 6 (Mb)
←HLA-DRB1
HLA-DQA1→
←HLA-DQB1
HLA-DQA2→
←HLA-DQB2

**B *NFKBIL1***

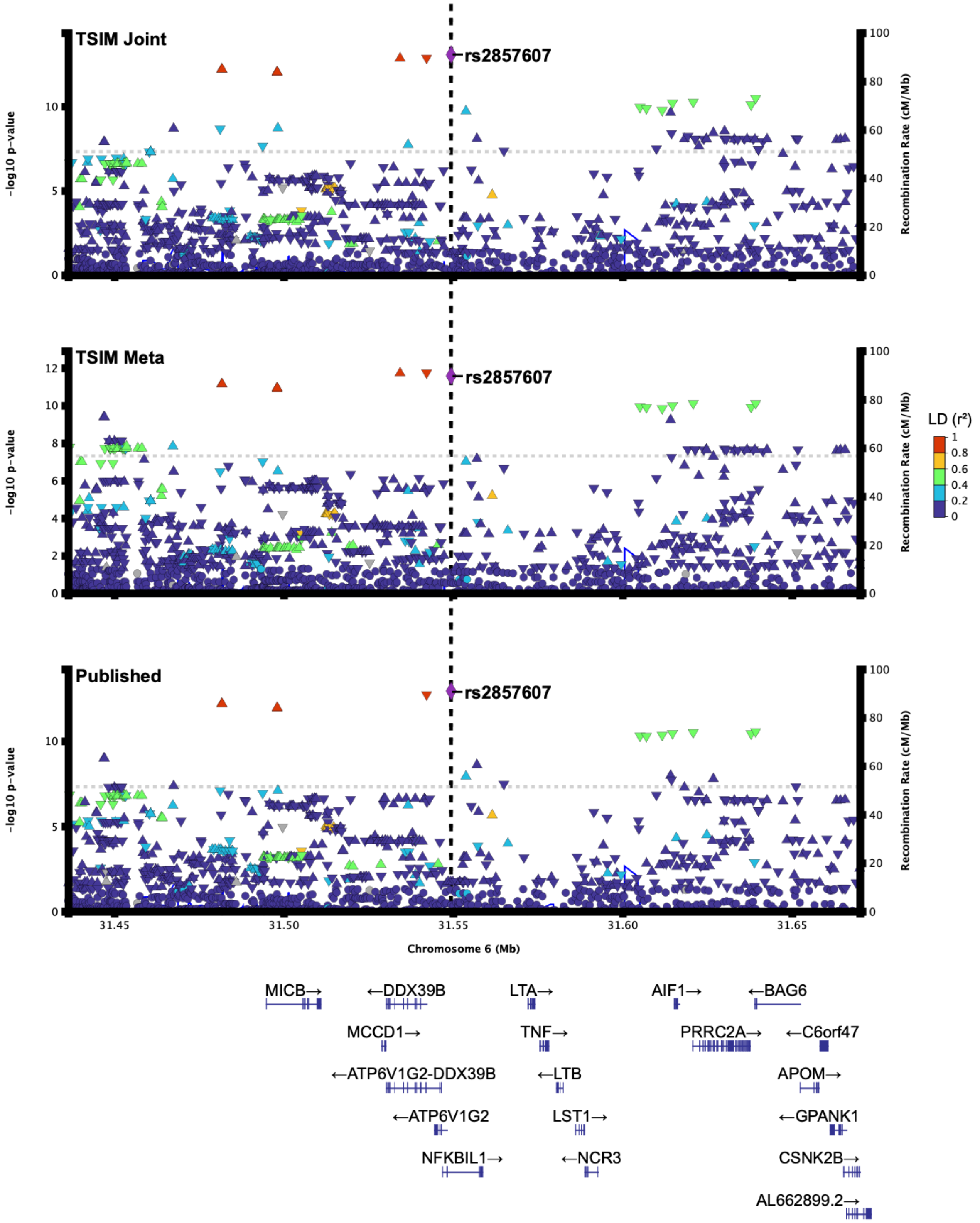
TSIM Joint
TSIM Meta
Published
rs2857607
-log10 p-value
Recombination Rate (cM/Mb)
LD (r²)
1
0.8
0.6
0.4
0.2
0
Chromosome 6 (Mb)
31.45
31.50
31.55
31.60
31.65
MICB→
←DDX39B
LTA→
AIF1→
←BAG6
MCCD1→
TNF→
PRRC2A→
←C6orf47
←ATP6V1G2-DDX39B
←LTB
APOM→
←ATP6V1G2
LST1→
←GPANK1
NFKBIL1→
←NCR3
CSNK2B→
AL662899.2→

C *CALHM6*

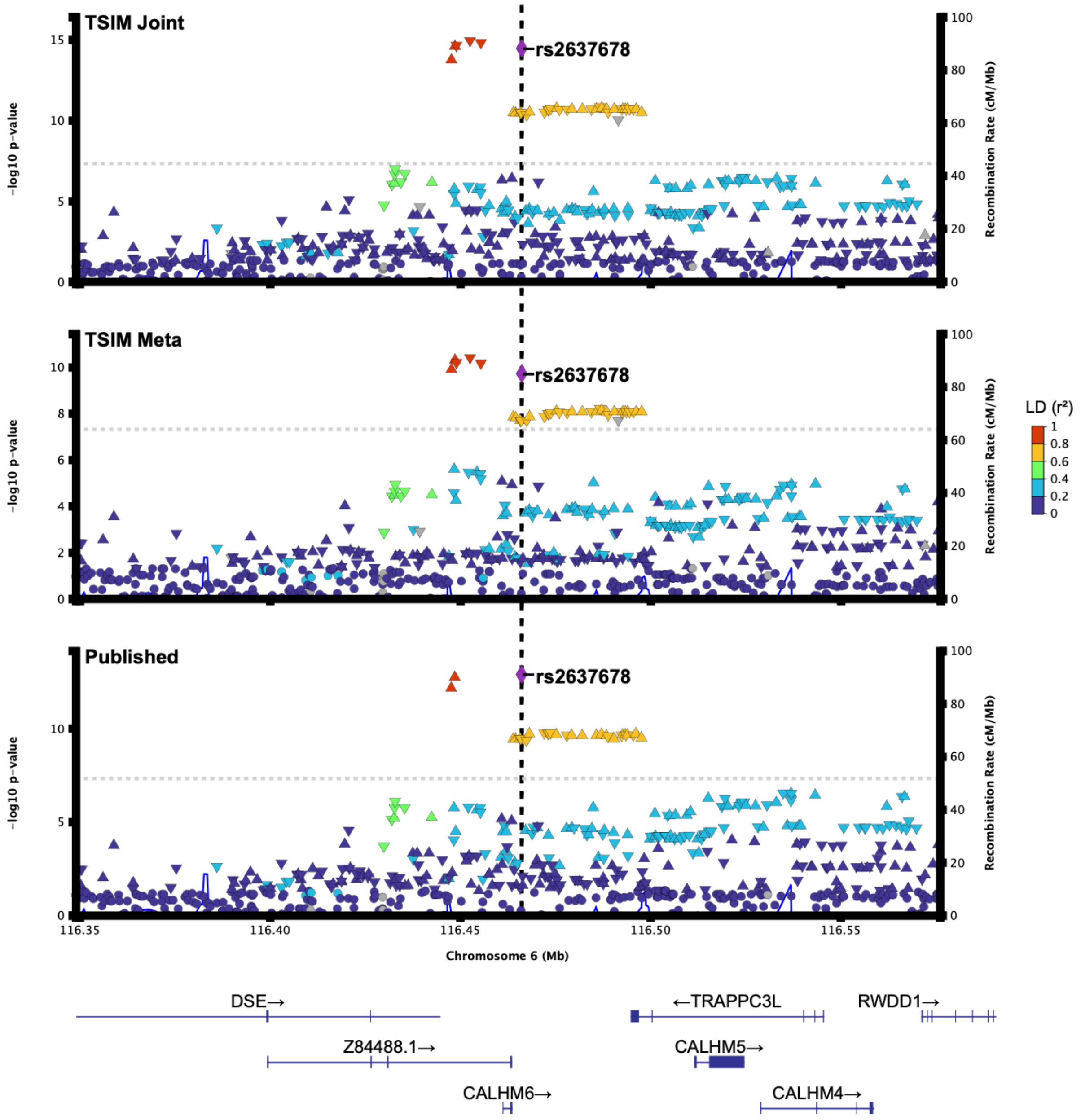
TSIM Joint
TSIM Meta
Published
rs2637678
-log10 p-value
Recombination Rate (cM/Mb)
LD (r²)
1
0.8
0.6
0.4
0.2
0
116.35
116.40
116.45
116.50
116.55
Chromosome 6 (Mb)
DSE→
Z84488.1→
CALHM6→
←TRAPPC3L
CALHM5→
CALHM4→
RWDD1→

**D *MORF4L***

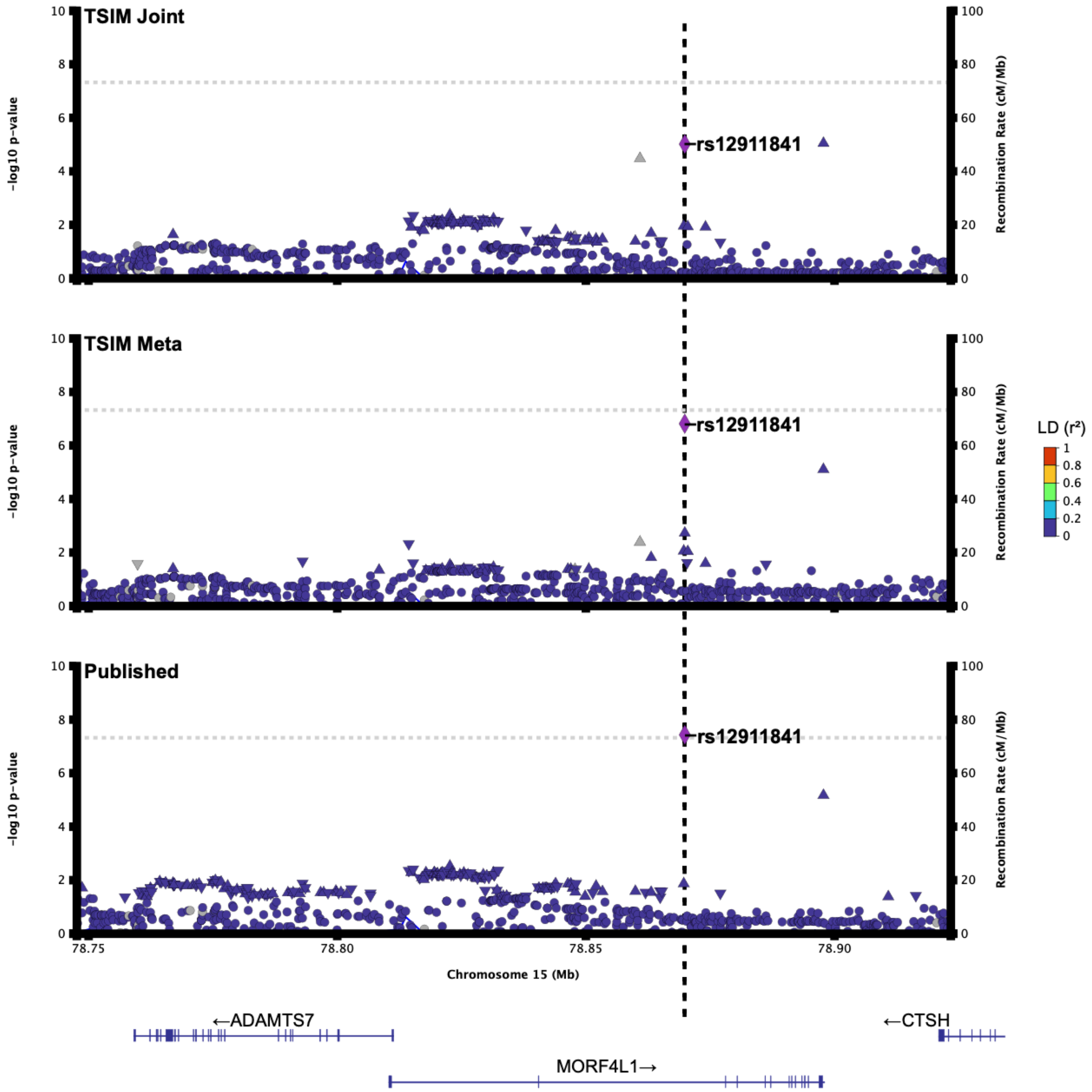


**Supplemental Figure S9: Locus plots for GWAS loci of pSSNS cohorts**

Locus plots highlighting SNPs detailed in **Supplemental Table S13** for *Published* GWAS (bottom), *TSIM-meta* (middle), and *TSIM-joint* (top) analyses. Gene loci include **(A)** *HLA-DQB1*, **(B)** *NFKBIL1*, **(C)** CALHM6, and **(D)** *MORF4L1*.

**Supplemental Table Legends**

**Supplemental Table S1: Correlation of imputed vs. WGS genotypes stratified by population and imputation quality (Imputed-only SNPs)**

Number of SNPs and correlation in each imputation quality bin and MAF bin shown in **Fig. 2A** for imputed-only SNPs.

**Supplemental Table S2: Correlation of imputed vs. WGS genotypes stratified by population and imputation quality (SNPs on Genotyping Array)**

Number of SNPs and correlation in each imputation quality bin and MAF bin shown in **Fig. 2B** for SNPs on genotyping array.

**Supplemental Table S3: Correlation of imputed vs. WGS genotypes stratified by population and empirical imputation quality (SNPs on Genotyping Array with $R^2$>0.98)**

Number of SNPs and correlation in each empirical imputation quality bin and MAF bin shown in **Fig. 2C** for SNPs on genotyping array. SNPs with $R^2$>0.98 were included only.

**Supplemental Table S4: Correlation of imputed vs. WES genotypes stratified by population and imputation quality (Imputed-only SNPs)**

Number of SNPs and correlation in each imputation quality bin and MAF bin shown in **Supplemental Fig. S1A** for imputed-only SNPs.

**Supplemental Table S5: Correlation of imputed vs. WES genotypes stratified by population and imputation quality (SNPs on Genotyping Array)**

Number of SNPs and correlation in each imputation quality bin and MAF bin shown in **Supplemental Fig. S1B** for SNPs on genotyping array.

**Supplemental Table S6: Correlation of imputed vs. WES genotypes stratified by population and empirical imputation quality (SNPs on Genotyping Array)**

Number of SNPs and correlation in each empirical imputation quality bin and MAF bin shown in **Supplemental Fig. S1C** for SNPs on genotyping array.

**Supplemental Table S7: Correlation of imputed vs. WGS genotypes by stage**

Number of SNPs and correlation in each imputation quality bin and MAF bin shown in **Fig. 3** and **Supplemental Fig. S2**.

**Supplemental Table S8: Correlation of imputed vs. WES genotypes by stage**

Number of SNPs and correlation in each imputation quality bin and MAF bin shown in **Supplemental Fig. S3.**

**Supplemental Table S9: Metadata and results of arthritis and UKB GWAS scenarios**

Metadata and results of arthritis and UKB GWAS scenarios. Columns: Scenario = indicates which datasets were merged, Cases = number of cases, Controls = number of controls, Total samples = total number of samples, PCs = number of PCs included as covariates, *Separately-imputed* GWAS = results of GWAS performed on cohorts merged after single, separate imputations, *TSIM* GWAS= results of GWAS performed on cohorts merged with TSIM, *Published* GWAS = results of GWAS performed on just arthritis dataset; SNPs = number of SNPs included in GWAS, $\lambda$ = genomic control calculated based on the median, GWAS loci = total number of independent genome-wide significant loci found.

**Supplemental Table S10: Detailed information on genome-wide significant loci for arthritis and UKB5K GWAS**

Genome-wide significant loci identified in either *Published* (arthritis cases and controls only), *TSIM external controls* GWAS (merged arthritis cases with UKB5K), or *TSIM both controls* (merged arthritis cases and controls with UKB5K). Columns: Loci ID = letter associating all variants attributed to the same loci as one (based on LD), Variant ID = variant chromosome, position (hg38), and ref/alt alleles, rsID in LD = rsID of variants in high LD with variant in rsID/Variant ID (all variants in high LD have the same loci ID), EA = effect allele, EAF = effect allele frequency, P-values = unadjusted p-values, $\lambda_{GC}$-Adj. P-value = p-values from GWAS adjusted by genomic control

($\lambda_{GC}$), OR = odds ratios, Imputation Quality = $R^2$ reported by Minimac for variant in rsID/VariantID, Genotyped? = whether the SNP was genotyped (i.e., present on genotyping array) for both stages of imputation.

**Supplemental Table S11: Cohort metadata for pSSNS GWAS**
Information about the cohorts used for pSSNS GWAS. Columns: Subcohort = subcohort label based on where dataset came from, Cohort = cohort that study belongs to, Genotyping platform = genotyping platform used to genotype dataset, Cases = number of cases, Controls = number controls, Total samples = total number of samples.

**Supplemental Table S12: Metadata for results of the pSSNS GWAS analyses**
Metadata for results of the pSSNS GWAS analyses. Columns: Analysis = label for analysis conducted, SNPs = number of SNPs included in analysis, PCs (EU/US) = number of PCs included as covariates (for meta-analyses, PCs used for EU and PCs used for US are indicated as EU/US), $\lambda$ = genomic control calculated based on the median; TSIM Joint = single GWAS conducted on all studies merged using TSIM, TSIM Meta = meta-analysis of GWAS conducted on EU, merged using TSIM, and US cohorts, Published Meta = results of meta-analysis from Barry et al. without Dufek et al. data, TSIM/Published EU = results from GWAS of EU cohort, TSIM/Published US = results from GWAS of US cohort.

**Supplemental Table S13: Detailed information on genome-wide significant loci for pSSNS GWAS analyses**
Detailed information on genome-wide significant loci for pSSNS GWAS analyses. Results from TSIM GWAS are in top subtable and results from Published GWAS without Dufek et al. are in bottom subtable. Columns: Variant ID = variant chromosome, position (hg38), and ref/alt alleles, EA = effect allele, EAF = effect allele frequency, P-values = unadjusted p-values, $\lambda_{GC}$-Adj. P-value = p-values from GWAS adjusted by genomic control ($\lambda_{GC}$), OR = odds ratios, Imputation Quality = $R^2$ reported by Minimac for variant in rsID/VariantID, Genotyped? = whether the SNP was "genotyped" (i.e., in the input VCF) for both stages of imputation; TSIM Joint = single GWAS conducted on all studies merged using TSIM, Meta = meta-analysis of GWAS conducted on EU and US cohorts, EU = results from GWAS of EU cohort, US = results from GWAS of US cohort.
